# Maternal depressive symptoms, neonatal white matter, and toddler social-emotional development

**DOI:** 10.1101/2022.04.28.22274095

**Authors:** Alexandra Lautarescu, Alexandra F. Bonthrone, Maximilian Pietsch, Dafnis Batalle, Lucilio Cordero-Grande, J-Donald Tournier, Daan Christiaens, Joseph V Hajnal, Andrew Chew, Shona Falconer, Chiara Nosarti, Suresh Victor, Michael C. Craig, A. David Edwards, Serena J. Counsell

**Affiliations:** Centre for the Developing Brain, Department of Perinatal Imaging and Health, School of Biomedical Engineering and Imaging Sciences, King’s College London, St Thomas’ Hospital, London, United Kingdom; Department of Psychology, Institute of Psychiatry, Psychology and Neuroscience, King’s College London, London, United Kingdom; Department of Child and Adolescent Psychiatry, Institute of Psychiatry, Psychology and Neuroscience, King’s College London, London, United Kingdom; National Female Hormone Clinic, South London and Maudsley National Health Service Foundation Trust, London, United Kingdom; Biomedical Research Networking Center in Bioengineering, Biomaterials and Nanomedicine (CIBER-BBN), Madrid, Spain; Biomedical Image Technologies, ETSI Telecomunicación, Universidad Politécnica de Madrid, Madrid, Spain; Department of Electrical Engineering, ESAT/PSI, KU Leuven, Leuven, Belgium; MRC Centre for Neurodevelopmental Disorders, King’s College London, London, United Kingdom; EPSRC/Wellcome Centre for Medical Engineering, King’s College London, London, United Kingdom; Neonatal Unit, Evelina London Children’s Hospital, London, United Kingdom

## Abstract

Maternal prenatal depression is associated with increased likelihood of neurodevelopmental and psychiatric conditions in offspring. The relationship between maternal depression and offspring outcome may be mediated by in-utero changes in brain development. Recent advances in magnetic resonance imaging (MRI) have enabled in vivo investigations of neonatal brains, minimising the effect of postnatal influences. The aim of this study was to examine associations between maternal prenatal depressive symptoms, infant white matter, and toddler behaviour. 413 mother-infant dyads enrolled in the developing Human Connectome Project. Mothers completed the Edinburgh Postnatal Depression Scale (median = 5, range = 0-28, n=52 scores ≥ 11). Infants (n=223 male) (median gestational age at birth=40 weeks, range 32.14-42.29) underwent MRI (median postmenstrual age at scan=41.29 weeks, range 36.57-44.71). Fixel-based fibre metrics (mean fibre density, fibre cross-section, and fibre density modulated by cross-section) were calculated from diffusion imaging data in the left and right uncinate fasciculi and cingulum bundle. For n=311, internalizing and externalizing behaviour, and social-emotional abilities were reported at a median corrected age of 18 months (range 17-24). Statistical analysis used multiple linear regression and mediation analysis with bootstrapping. Maternal depressive symptoms were positively associated with infant fibre density in the left (B =.0005, p=.003, q=.027) and right (B=.0006, p=.003, q=.027) uncinate fasciculus, with the left uncinate fasciculus, in turn, positively associated with social-emotional abilities in toddlerhood (B =105.70, p=.0007, q=.004). In a mediation analysis, higher maternal depressive symptoms predicted toddler social-emotional difficulties (B=.342, t(307)=3.003, p=.003), but this relationship was not mediated by fibre density in the left uncinate fasciculus (Sobel test p=.143, bootstrapped indirect effect=.035, SE=.02, 95%CI [-.01,.08]). There was no evidence of an association between maternal depressive and cingulum fibre properties. These findings suggest that maternal perinatal depressive symptoms are associated with neonatal uncinate fasciculi microstructure, but not fibre bundle size, and toddler behaviour.

## 1. Introduction

Maternal depression is one of the most common prenatal complications and is related to poor neurodevelopmental and psychiatric outcomes in offspring (1). These outcomes may be explained, at least in part, by changes in the developing brain during intrauterine life (2, 3). However, most research is based on older participants, making it difficult to separate the effects of prenatal and postnatal environment.

Recent advances in magnetic resonance imaging (MRI) have enabled in vivo investigations of neonatal brains (4). A growing number of diffusion MRI studies suggest that maternal depression and anxiety are associated with changes in the infant brain, with reports of lower Fractional Anisotropy (FA) (5–7) and higher FA (8, 9) in widespread regions. Previous literature suggests that frontal-limbic areas including the uncinate fasciculus and the cingulum may be particularly vulnerable to the effects of maternal stress (10–15).

Changes in the micro- and macrostructure of these tracts have also been linked to the development of mood disorders, social information processing, and behavioural and social development (16–20).

Neonatal microstructure of the right anterior cingulum bundle is associated with increased internalizing symptoms, inattention and impaired social cognition at 5 in children born prematurely (21). Altered white matter microstructure (22), particularly in the uncinate fasciculus has been linked with social difficulties in young autistic children (19), and emotional development in children born very preterm (23). Altered white matter development in the neonatal period may be an antecedent for social communication or behavioural impairments in early childhood.

It is important to note that outcomes in offspring exposed prenatally to poor maternal psychological wellbeing are characterised by a large degree of heterogeneity, and not all children have poor outcomes (1, 2). There is also evidence that male and female offspring may be affected differently (24). Similarly, adverse outcomes and altered brain structure and function are not just associated with maternal clinical depression, but also with subthreshold depressive symptoms (2, 3), emphasizing the importance of using a dimensional approach.

It is possible that individual differences in developmental outcomes in at risk infants are in part explained by differences in early brain development. Therefore, it is necessary to understand whether the association between maternal depressive symptoms and behavioural outcomes is mediated by early brain development. Prior mediation and moderation studies have mainly focused on biological indicators of stress (25, 26), with only a few studies investigating self-reported psychological wellbeing (27, 28). Moog and colleagues (28) reported that variations in hippocampal volume may mediate the association between maternal perceived stress and infant social-emotional function.

The diffusion MRI literature linking maternal self-reported depressive symptoms with offspring brain development and behaviour is scarce. Hay and colleagues (13) reported that amygdala-prefrontal structural connectivity mediated the association between prenatal depression and externalizing behaviour in males. However, imaging in this study was performed at 4 years of age, making it difficult to separate the effects of the prenatal and postnatal environment. To our knowledge, there has been only one such study in a smaller neonatal sample (n=80, (15)) which reported that, while there was no direct association between maternal depressive symptoms and white matter properties, whole-brain FA moderated the association between maternal depression and negative reactivity.

It is important to note that the majority of these studies are based on the diffusion tensor model (29) which is limited by its inability to describe more than one dominant fibre orientation and to provide accurate information in voxels that contain crossing and kissing fibres (30). However, crossing fibres exist in over 90% of white matter voxels in the adult brain (31). In addition, DTI metrics may be affected by multiple features of the underlying microstructure.

Modern data acquisition techniques such as those employed as part of the developing Human Connectome Project (dHCP, developingconnectome.org) allow more information to be extracted from the diffusion-weighted signal. Fixel-based analysis (FBA, (32)) achieves microstructural and spatial specificity and characterises multiple fibre populations per voxel. FBA provides information regarding white matter structure for fibre bundles within a voxel through estimates of microstructure via local fibre density (FD), and morphometry via the fibre cross-section (FC) metric; their product, the fibre density and cross section (FDC), serves as a proxy of a fibre bundle’s capacity to relay information (32). This is of particular use in the neonatal brain where different white matter fibre populations within a voxel may be at different stages of development (33–35). Given its key advantages over DTI, the FBA framework is increasingly adopted in studies of both typical and clinical populations (see review by Dhollander and colleagues (36).

The primary aim of the current study was to improve our understanding of the relationship between maternal depressive symptoms and developing white matter using advanced neuroimaging in the largest neonatal sample to date.

We hypothesized that higher maternal depressive symptoms (as measured during the prenatal or early postnatal period) are associated with differences in FD, FC, and FDC in the uncinate fasciculus and cingulum in the neonatal brain. This study expands upon previous work from our group (11), shifting the focus towards depression and expanding the research question to also include the cingulum. A hypothesis-driven tract-specific approach was employed, in line with previous fixel-based work (see (36)); a direction of effect was not specified due to lack of comparable measures and inconsistencies in the literature to date.

Our secondary aim was to investigate the relationship between maternal depressive symptoms, neonatal white matter, and child behaviour assessed at 18 months of age, using a similar mediation analysis strategy as that of Hay and colleagues (13). We aimed to build on our recent work (37) which showed, in an overlapping sample, that higher maternal depressive symptoms were associated with toddlers’ higher internalizing and externalizing behaviour and social-emotional difficulties. We hypothesized that tracts with significant relationships with maternal depressive symptoms are associated with childhood internalizing and externalizing behaviour, and social-emotional difficulties. Thirdly, we hypothesized that associations between maternal depressive symptoms and behavioural outcomes are mediated by white matter micro- and/or macrostructure.

To our knowledge, this is the first study using fixel-based fibre metrics to investigate the relationship between maternal depressive symptoms and neonatal white matter development, as well as the first study to include multi-shell MR data allowing us to perform both fixel-based and DTI analyses.

## 2. Materials and methods

### 2.1. Participants

Infants were recruited between March 2015 and March 2021 as part of the dHCP. This study was approved by the UK National Research Ethics Authority (London - Riverside: 14/LO/1169) and conducted in accordance with the World Medical Association Code of Ethics (Declaration of Helsinki). Written informed parental consent was provided for all participants prior to data collection. Participants were invited to take part in one or more of the following: fetal MRI scan(s), neonatal MRI scan(s), and neurodevelopmental assessment. The analysis reported in this study is based on neonatal diffusion MRI (dMRI) data which, at the time the analysis was conducted, comprised data for n=894 scans. Scans were excluded for the following reasons: repeat scans (n=106), absence of completed Edinburgh Postnatal Depression Scale (EPDS, n=100), not singleton pregnancy (n=87), missing or incomplete dMRI data (n=85), failed dHCP image quality control (https://biomedia.github.io/dHCP-release-notes/qc.html) (n=21), gestational age (GA) at birth under 32 weeks (n=46), postmenstrual age (PMA) at scan under 37 weeks (n=11), major incidental findings as determined by a paediatric neuroradiologist (n=22, e.g., arterial ischemic infarcts, brain size <1^st^ centile), failed visual image quality control (n=2), no successful T2-weighted scan (n=1). The final sample for the MRI analysis included n=413 mother infant-dyads. A subset of n=311 participants also participated in a neurodevelopmental assessment conducted between 17-24 months post-expected delivery date.

### 2.2. Maternal and infant characteristics

Maternal depressive symptoms were measured using the EPDS (38) at each study visit. The EPDS is a 10-item screening tool (total score range 0-30). A cut-off of 11 or more has been shown to maximise the sensitivity and specificity of screening for depressive symptoms (39), while a score of 13 or more can be used to indicate high-level symptoms. EPDS scores were included in the analysis if completed during pregnancy (i.e., at a visit for a fetal scan) or in the early postnatal period (i.e., at the time of the first neonatal scan), as these scores were more likely to be reflective of mood during the prenatal and early postnatal period. EPDS scores in our sample were available as follows: n=4 only prenatal timepoints, n=289 only postnatal timepoint, n=120 both prenatal and postnatal timepoints. As the EPDS only asks about symptoms over the last 7 days, in cases where participants completed more than one EPDS questionnaire, the highest score was chosen. EPDS scores used in this analysis were collected at prenatal timepoints for n=55 (median GA=28.78 weeks, range 22.00-39.57) and first postnatal timepoint for n=358 (median GA=41.14 weeks, range 36.57-44.71). EPDS-3A scores were also calculated and reported, with a threshold of 6 or more suggesting anxiety symptoms (40).

Maternal history of poor mental health (coded as a binary yes/no) was determined based on multiple sources including maternal self-report, maternity notes, and mental health records from South London and Maudsley NHS Foundation Trust (see Supplement).

Sociodemographic characteristics and medical history for the mother-infant dyads were collected at enrolment (Table 1). As a proxy of socioeconomic status, the Index of Multiple Deprivation (IMD) was calculated from maternal postcode based on the 2019 IMD classification (41) with higher scores representing greater social deprivation. The relationship between infant GA at birth and PMA at scan is depicted in Fig S7.

**Table 1.**
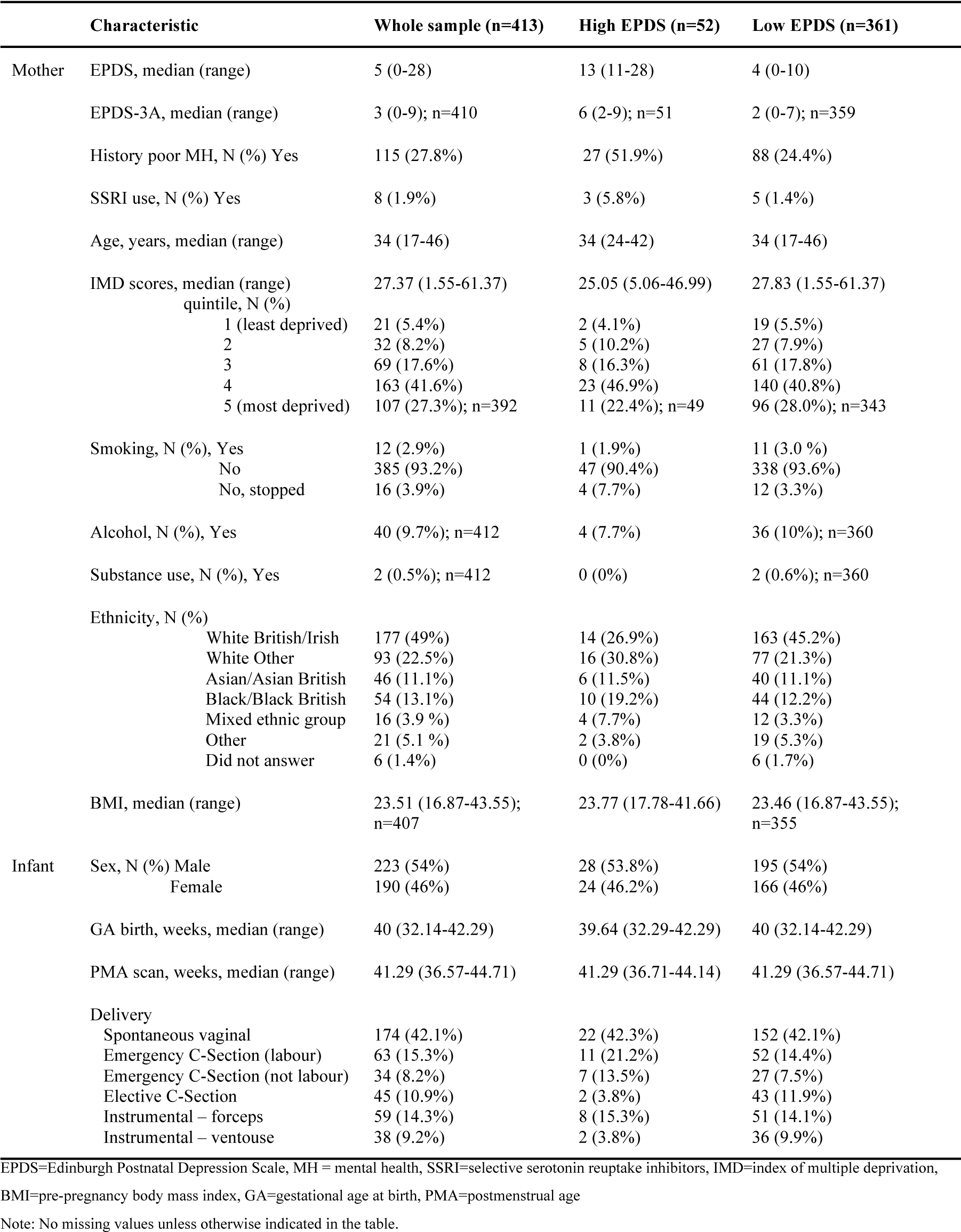
Maternal and infant characteristics

### 2.3. MR Imaging

#### 2.3.1. Acquisition

High resolution MRI of the neonatal brain was acquired on a Philips Achieva 3T system (Best, The Netherlands) using a dedicated 32-channel neonatal head coil and a neonatal positioning device (42). Neonates were scanned during natural sleep and monitored throughout the scan (see Supplement) whilst a paediatrician with experience in MRI was present.

The full dHCP protocol includes structural, functional, and diffusion imaging, but the current study is focused on the latter. Multi-shell High Angular Resolution Diffusion Imaging was acquired over 20 minutes using a protocol optimized for the neonatal brain (43, 44). For all completed scans, each dataset contained 300 volumes of Echo Planar Imaging slices, sampled with four phase-encode directions on four shells with b-values of 0 (n=20), 400 s/mm^2^ (n=64), 1000 s/mm^2^ (n=88) and 2600 s/mm^2^ (n=128). Acceleration of multiband 4, SENSE factor 1.2, and partial Fourier 0.86 were used, with acquired resolution 1.5 × 1.5 mm, 3 mm slices with 1.5 mm overlap, and TR/TE of 3800/90 ms. Images were processed with denoising (45), Gibbs ringing suppression (46), motion and image distortion correction using slice-to-volume reconstruction to a 1.5mm isotropic resolution (47), and inter-slice intensity inconsistency correction (48).

#### 2.3.2. Pre-processing

The processing pipeline of the dMRI data follows that of the FBA framework but was adapted to account for the properties of neonatal tissue (34). For all subjects, tissue-specific signal fingerprints (response functions) were estimated for single-fibre white matter and cerebrospinal fluid (CSF) using the Dhollander method (49), with an FA threshold of 0.15 for white matter and masks that exclude areas of CSF flow.

For group-analysis, response functions representative of free fluid and white matter diffusion signal at 44 weeks were calculated by the averaging all CSF response functions and 21 white matter response functions from subjects aged 44.1 (SD 0.3) weeks. These group-average response functions were used to calculate maps of free water density and tissue Fibre Orientation Distributions (FODs) using multi-shell multi-tissue constrained spherical deconvolution (50, 51). The estimated FODs were intensity normalised (52, 53) and conservative brain masks generated by intersecting masks generated in MRtrix3 (54) and by the brain extraction tool in FSL (55).

For group-analysis, a representative template was generated from the average of all subjects’ normalised white matter FOD and free water density images transformed from native to the “Schuh dHCP extended atlas” 40-week anatomical template space (56). These transformations are composite warps consisting of transformations released as part of the dHCP data release 3: a rigid-body subject- dMRI to subject-T2w (estimated with FSL flirt using a normalised mutual information metric), subject-T2w to age-matched T2-weighted template (estimated with nonlinear diffeomorphic multimodal registration of T2-weighted image and grey matter/white matter (GM/WM) tissue probability maps using the diffeomorphic symmetric image normalization method (SyN) in the Advanced Normalization Tools (ANTs) software package (57)) and week-to-week nonlinear transformations to the 40-week dHCP extended atlas template (estimated using the template T2- weighted images and SyN registration). The aggregate warps were estimated on a 1.3 isotropic grid, yielding a study-specific two-tissue template with the same resolution.

Each subject’s normalised white matter FOD and fluid density images were jointly coregistered to the study specific template based on (58) using a normalised cross correlation metric, maximum spherical harmonics order lmax=2 and a multi-resolution pyramid with 66 stages from 3.75mm to 1.5mm voxel size. FODs and masks were warped to the template space using cubic and linear interpolation, respectively.

All T2-weighted images were motion corrected and reconstructed to a 0.8 mm isotropic resolution (59), images were processed and segmented using the dHCP structural pipeline (60, 61) (see Supplement).

#### 2.3.3. Tractography in template space

Tractography of the left and right uncinate fasciculus, dorsal cingulum and ventral cingulum was performed on the white matter template using an anatomically constrained tractography (ACT) probabilistic algorithm with 50,000,000 seeds per tract (Fig 1). ACT (62) was performed in template space using the draw-EM parcellation of the dHCP 40-week extended atlas template.

**Fig 1.**
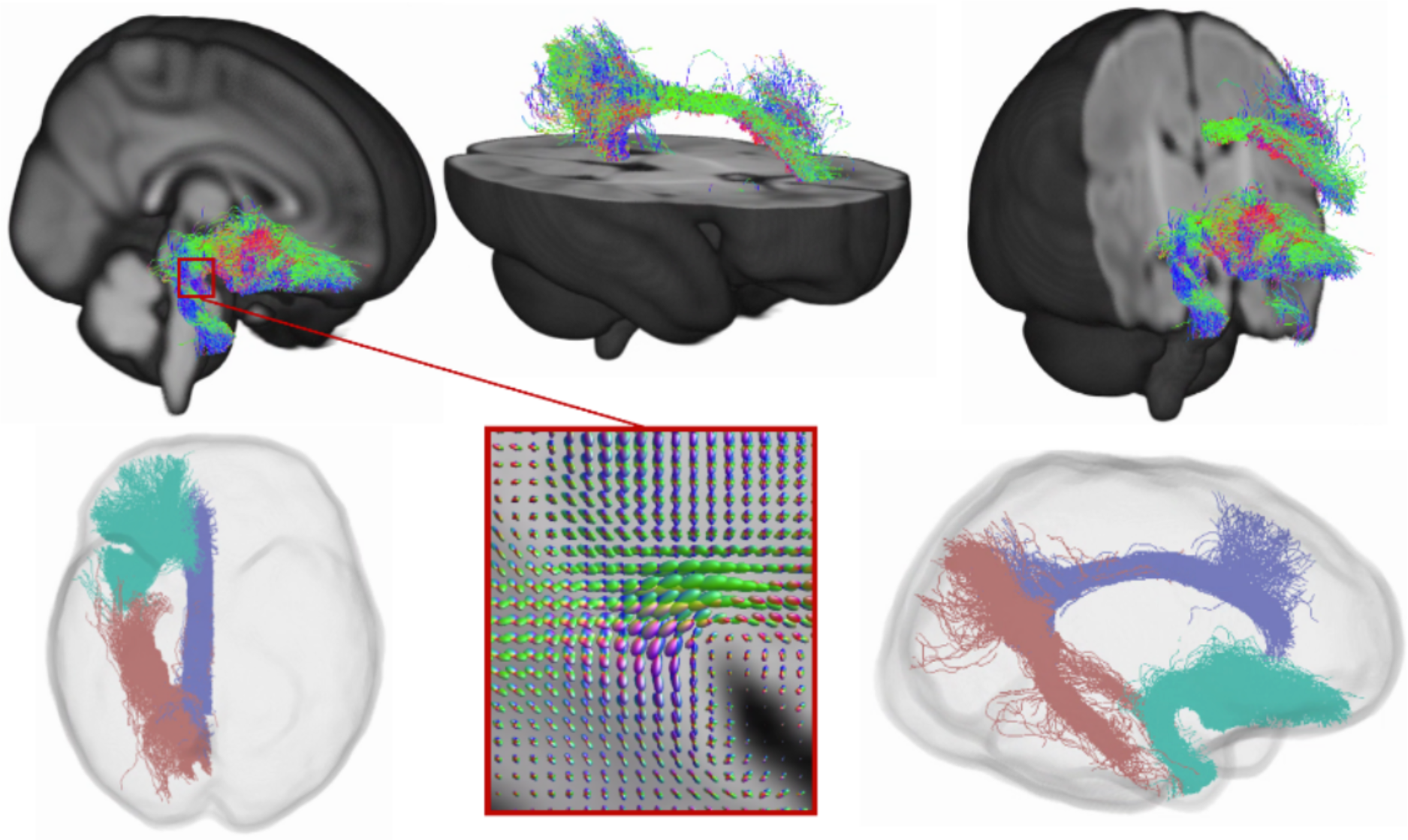
Visual representation of the tracts in template space. The top row shows the location of the tracts, coloured by streamline orientation (blue: superior-inferior, red: left-right, green: anterior-posterior). On the bottom row, the middle image shows an example of fibre orientation distributions in a region of crossing fibres, while the left and right images represent “glass-brain” illustrations of the tracts of interest (pink: ventral cingulum, purple: dorsal cingulum, turquoise: uncinate fasciculus).

Regions of interest for tractography of the left and right uncinate fasciculus and dorsal and ventral cingulum (Table S1) were obtained from a neonatal version (63) of the anatomical automatic labelling (AAL) atlas (56, 64) adapted to the dHCP template. This was registered to the dHCP 40-week extended atlas template space using ANTs SyN (57).

#### 2.3.4. Fixel-based fibre metrics

Tracts in template space were converted to fixel masks and mean FD, FC, and FDC were extracted across each tract for each baby, according to the MRtrix3 pipeline. Measures were averaged over all fixels. Apparent FD is, at high diffusion-weightings and subject to certain conditions (65), approximately proportional to the volume of the intra-axonal compartment. Sensitivity of apparent fibre density (AFD) to the tissue microstructure of individual fibre populations within a voxel (“fixels”) can be achieved by segmenting the FOD lobes (66) and numerical integration over directions corresponding to the fixels. FC is a local measure of the change of a fixel’s cross-sectional area due to warping to the joint template space (i.e., local expansion and contraction perpendicular to the bundle), with increases in this metric being thought to reflect an increase in the cross-sectional spatial extent occupied by a tract. FC was calculated from the subject-to-template warps estimated with mrregister and log-transformed (log(FC)) in accordance with the MRtrix3 pipeline recommendations. FDC is a composite metric reflecting both microstructural and morphological differences; it was calculated by multiplying FD and FC (67, 68)

#### 2.3.5. Diffusion tensor imaging

To complement the information extracted from the FBA and enable comparison with previous literature, we undertook a secondary analysis using diffusion tensor imaging. FA and mean diffusivity (MD) maps for each infant were calculated from the b=0 and b=1000 shells of the dMRI data using MRtrix3. Tracts were warped from template space into native space for each infant, and mean DTI metrics for each tract in each infant were extracted and used for statistical analysis.

### 2.4. Child characteristics

A neurodevelopmental assessment was conducted at approximately 18 months of age. Outcomes of interest for the current study included parent-reported behavioural outcomes measured using the Child Behaviour Checklist (CBCL) for ages 1.5 to 5 (69) and social-emotional abilities measured using the Quantitative Checklist for Autism in Toddlers (Q-CHAT, (70). The CBCL includes 100 items and subscales for internalizing behaviours (i.e., being withdrawn, somatic complaints, anxiety, depression, emotional reactivity) and externalizing behaviours (i.e., attention problems, aggressive behaviours).

The Q-CHAT includes 25 items evaluating social communication, as well as repetitive, stereotyped, and sensory behaviours. Cognitive development was assessed using the Bayley Scales of Infant and Toddler Development, 3^rd^ Edition, 2006 (BSID-III, (71)), home environment was assessed with the Stimulating Parenting Scale (72) and dysfunctional parenting was assessed with the Parenting Scale (73) (Table 2).

**Table 2.** Behavioural and cognitive outcomes in toddlers

| Characteristic | Whole sample (n=311) | High EPDS (n=35) | Low EPDS (n=276) |
| --- | --- | --- | --- |
| IMD 18 months, median (range) | 25.26 (1.55-61.37); n=292 | 25.05 (5.06-53.18); n=33 | 25.26(1.55-61.37); n=259 |
| quintile, N (%) |  |  |  |
| 1 (least deprived) | 28 (9.6%) | 2 (6.1%) | 26 (10.0%) |
| 2 | 32 (11.0%) | 3 (9.1%) | 29 (11.2%) |
| 3 | 52 (17.8%) | 4 (12.1%) | 48 (18.5%) |
| 4 | 110 (37.7%) | 15 (45.5%) | 95 (36.7%) |
| 5 (most deprived) | 70 (24.0%) | 9 (27.3%) | 61 (23.6%) |
| Corrected age, months, median (range) | 18 (17-24) | 18 (17-24) | 18 (17-24) |
| CBCL total T-score, median (range) | 46 (28-69) | 52 (37-66) | 46 (28-69) |
| N (%), Normal ($\leq 64$) | 301 (96.8%) | 32 (91.4%) | 269 (97.5%) |
| Borderline | 10 (3.2%) | 3 (8.6%) | 7 (2.5%) |
| Clinical ($\geq 70$) | 0 (0%) | 0 (0%) | 0 (0%) |
| CBCL Int. T-score, median (range) | 43 (29-72) | 49 (29-70) | 43 (29-72) |
| CBCL Ext. T-score, median (range) | 48 (28-70) | 54 (28-65) | 47 (28-70) |
| Q-CHAT Total, median (range) | 29 (8-59) | 32 (20-52) | 28 (8-59) |
| N (%), Normal | 272 (87.5%) | 27 (77.1%) | 245 (88.8%) |
| High ($\geq 39$) | 39 (12.5%) | 8 (22.9%) | 31 (11.2%) |
| BSID-III Cog. Comp., median (range) | 100 (70-125) | 100 (70-120) | 100 (70-125) |
| N (%), Normal ($>85$) | 291 (93.6%) | 32 (91.4%) | 259 (93.8%) |
| Mild | 20 (6.4%) | 3 (8.6%) | 17 (6.2%) |
| Mod/severe ($<70$) | 0 (0%) | 0 (0%) | 0 (0%) |
| Home environment, median (range) | 21 (7-28); n=305 | 20 (12-28) | 21 (7-28); n=270 |
| Dysfunctional parenting, median (range) | 2.90 (1.43-4.20); n=308 | 3.20 (2.37-4) | 2.83 (1.43-4.20); n=273 |

EPDS=Edinburgh Postnatal Depression Scale, IMD=index of multiple deprivation, CBCL=Childhood Behaviour Checklist, Q-CHAT=Quantitative Checklist for Autism in Toddlers, Int=internalising, Ext=externalising, BSID-III Cog Comp = Bayley's cognitive composite score. Home environment = score on the Stimulating Parenting Scale, Dysfunctional parenting = score on the Parenting Scale.
Note: No missing values unless otherwise indicated in the table. The cut-offs used to indicate high scores on the CBCL, Q-CHAT, and BSID-III as are per (82-84, 132) and are used for descriptive purposes only.

### 2.5. Statistical analysis

Statistical analysis was performed using R version 4.1.1. (https://www.r-project.org/).

#### 2.5.1. Regression

Multiple linear regressions were used to test the relationship between EPDS and brain measures, and the relationship between brain measures and child outcomes. Covariates were chosen a priori based on theoretical considerations and associations with variables of interest (See Supplement).

First, using multiple linear regression, infant fixel-based fibre metrics were tested separately as dependent variables, with maternal EPDS and an EPDS-sex interaction as predictors. If the EPDS-sex interaction was not significant, it was excluded from the model and the analysis was rerun (as per (13)). The following covariates were also included in the models: GA at birth, PMA at scan, infant sex, maternal IMD at enrolment, maternal history of mental health (See Supplement). For FDC and log(FC) models, ICV was also included as a covariate due to known associations with these metrics (e.g., (68)). Correction for multiple comparisons was performed using the False Discovery Rate (FDR) based on 6 tracts (i.e., left and right uncinate fasciculus, dorsal cingulum, ventral cingulum) and 3 values each (mean FD, log(FC), FDC), for a total of 18 comparisons.

Secondly, tracts with significant (after FDR correction) associations with maternal EPDS scores were analysed for associations with toddler behaviour. CBCL scores (internalising and externalising T scores) and Q-CHAT scores (total score) were tested separately as dependent variables, with the FBA metric and an FBA-sex interaction as predictors. If the FBA-sex interaction was not significant, it was excluded from the model and the analysis was rerun. The following covariates were also included in the models: infant sex, infant GA at birth, infant PMA at scan, maternal IMD at the 18-month assessment, child corrected age at assessment, BDSID-III cognitive score, and maternal age (see Supplement).

Coefficients of determination (R^2^) and F statistics (F) are reported for each regression model. Unstandardised coefficients (B), t-values (t) and uncorrected p-values (p) are reported for the EPDS or EPDS by sex variables. FDR-corrected (q) p-values are reported for all significant uncorrected p-values (p<0.05).

Assumptions for regression were checked (linearity, homoscedasticity, independence, normality). Where assumptions for multiple regression were not met (see Supplement), robust regression was performed using lmrob from the robustbase R package, which uses fast-S algorithms and heteroscedasticity and autocorrelation corrected (HAC) standard errors (74). Robust regression is designed to be resistant to outlying observations and non-normality (75).

#### 2.5.2. Mediation

A mediation analysis was conducted to investigate whether the relationship between maternal EPDS scores and toddler behaviour is mediated by changes in neonatal white matter. FBA metrics that were significantly associated with both EPDS scores and behaviour were selected for mediation analysis, with maternal EPDS scores as the predictor (X), FBA metrics as the mediator (M), and behaviour scores as the outcome variable (Y). Child age at neonatal scan and age at neurodevelopmental assessment were entered as covariates. Mediation was performed with the MeMoBootR package in R (76), with 5000 bootstrapped samples. This has been argued to be more powerful than the original causal steps mediation approach (77).

## 3. Results

Mean values for the FBA and the DTI metrics for each tract are provided in Table S2 and S3. For all tracts, FA was positively associated with FD (rho between .90 and .97, p<.001) and FDC (rho between .58 and .77, p<.001), and MD was negatively associated with FD (rho between -.87 and -.93, p<.001) and FDC (rho between -.50 and -.63, p<.001); both showed weak associations with log(FC) (Table S4).

### 3.1. Maternal depressive symptoms and neonatal white matter

Assumptions for multiple regressions were met for all models unless otherwise specified (See Supplement). Results are summarised in Table 3.

**Table 3.** Associations between maternal depressive symptoms and infant white matter

|  |  | EPDS model |  |  | EPDS x Sex (female) model |  |  | Model fit |  |
| --- | --- | --- | --- | --- | --- | --- | --- | --- | --- |
| Tract | Metric | B | t | p (q) | B | t | p (q) | R<sup>2</sup> | F(df) |
| UF-L | FD | .0005 | 2.96 | .003 (.027)* |  |  |  | .58 | F<sub>(6,385)</sub>=90.11 |
|  | FDC |  |  |  | -.0008 | -2.62 | .009 (.054) | .73 | F<sub>(8,383)</sub>=134.1 |
|  | Log(FC) | -.0007 | -1.33 | .183 |  |  |  | .72 | F<sub>(7,384)</sub>=143.4 |
| UF-R | FD | .0006 | 3.04 | .003 (.027)* |  |  |  | .57 | F<sub>(6,385)</sub>=87.92 |
|  | FDC | .0004 | 2.30 | .022 (.099) |  |  |  | .74 | F<sub>(7,384)</sub>=158.3 |
|  | Log(FC) | -.0007 | -1.37 | .171 |  |  |  | .74 | F<sub>(7,384)</sub>=157.6 |
| CD-L | FD | .0003 | 1.66 | .099 |  |  |  | .34 | F<sub>(6,385)</sub>=35.1 |
|  | FDC | .0003 | 2.10 | .037 (.133) |  |  |  | .65 | F<sub>(7,384)</sub>=102.8 |
|  | Log(FC) | .0003 | 0.53 | .592 |  |  |  | .78 | F<sub>(7,384)</sub>=201.2 |
| CD-R | FD | .0002 | 1.40 | .160 |  |  |  | .37 | F<sub>(6,385)</sub>=38.89 |
|  | FDC | .0003 | 1.71 | .088 |  |  |  | .66 | F<sub>(7,384)</sub>=110.6 |
|  | Log(FC) | .0003 | 0.52 | .607 |  |  |  | .74 | F<sub>(7,384)</sub>=157.9 |
| CV-L | FD | .0002 | 1.23 | .218 |  |  |  | .40 | F<sub>(6,385)</sub>=45.04 |
|  | FDC | .0002 | 1.10 | .271 |  |  |  | .66 | F<sub>(7,384)</sub>=110 |
|  | Log(FC) | -.0001 | -.32 | .750 |  |  |  | .80 | F<sub>(7,384)</sub>=223.9 |
| CV-R | FD | .0002 | 1.21 | .228 |  |  |  | .44 | F<sub>(6,385)</sub>=51.8 |
|  | FDC | .0001 | 0.91 | .362 |  |  |  | .68 | F<sub>(7,384)</sub>=120.3 |
|  | Log(FC) | -.00007 | -0.16 | .870 |  |  |  | .79 | F<sub>(7,384)</sub>=214 |

UF-L = left uncinate fasciculus, UF-R = right uncinate fasciculus, CD-L = left dorsal cingulum, CD-R = right dorsal cingulum, CV-L = left ventral cingulum, CV-R = right ventral cingulum, B=unstandardized coefficients, q=p values after FDR correction. \*Results that survive FDR correction for multiple comparisons. Note: All models (R squared) were significant at p<.001. The R squared values reported are the adjusted ones, given that models have different numbers of variables. Covariates were gestational age at birth, postmenstrual age at scan, infant sex, maternal IMD at enrolment, maternal history of mental health. In addition, for FDC and log(FC) models, ICV was also included as a covariate. If EPDS by sex interaction was not significant, it was removed from the model and these cells are blank.

#### 3.1.1. Uncinate fasciculus

EPDS scores had a positive main effect on left uncinate fasciculus FD (B =.0005, t(385)=2.958, p=.003) and right uncinate fasciculus FD (B=.0006, t(385)=3.036, p=.003), so that infants of mothers with higher EPDS scores had higher FD (Fig 2). Both relationships survived FDR correction for multiple comparisons (both q=.027), but the effect size was small compared to that of variables such as postmenstrual age at scan, and infant sex (Fig 3, Table S12).

**Fig 2.**
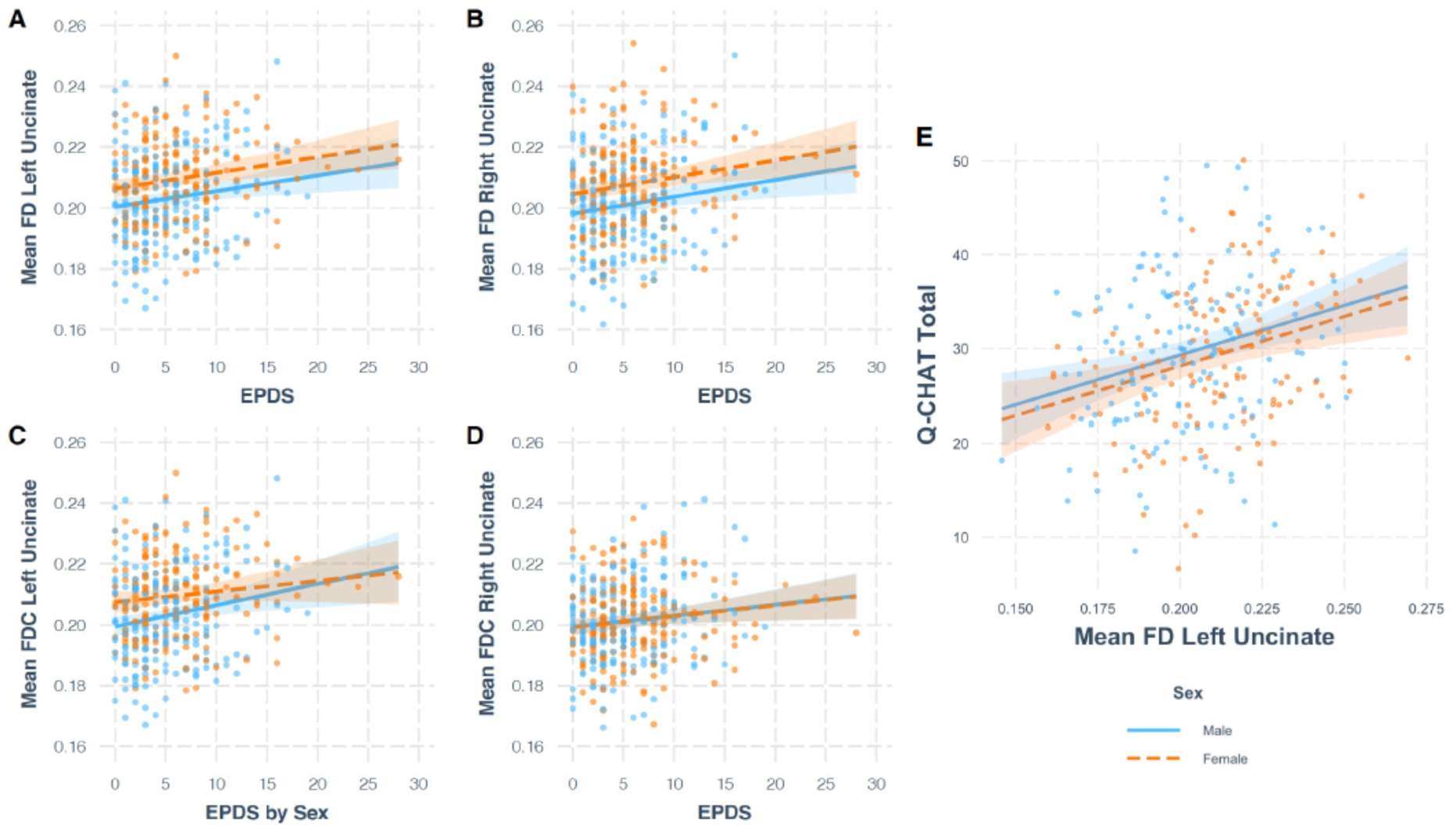
Panels A, B, C, D contain plots showing the relationship between EPDS and white matter microstructure and macrostructure, controlling for effect of covariates. For mean FD left (A) and right (B) uncinate fasciculus and mean FDC right uncinate fasciculus (D), the main model is plotted. For mean FDC left uncinate fasciculus (C), the model with EPDS by Sex interaction is plotted. The relationship between EPDS and mean FD in the left and right uncinate fasciculus (A, B) survived FDR correction. Panel E contains a plot from robust regression, showing the relationship between mean FD in the left uncinate fasciculus and total Q-CHAT score, controlling for the effect of covariates.

**Fig 3.**
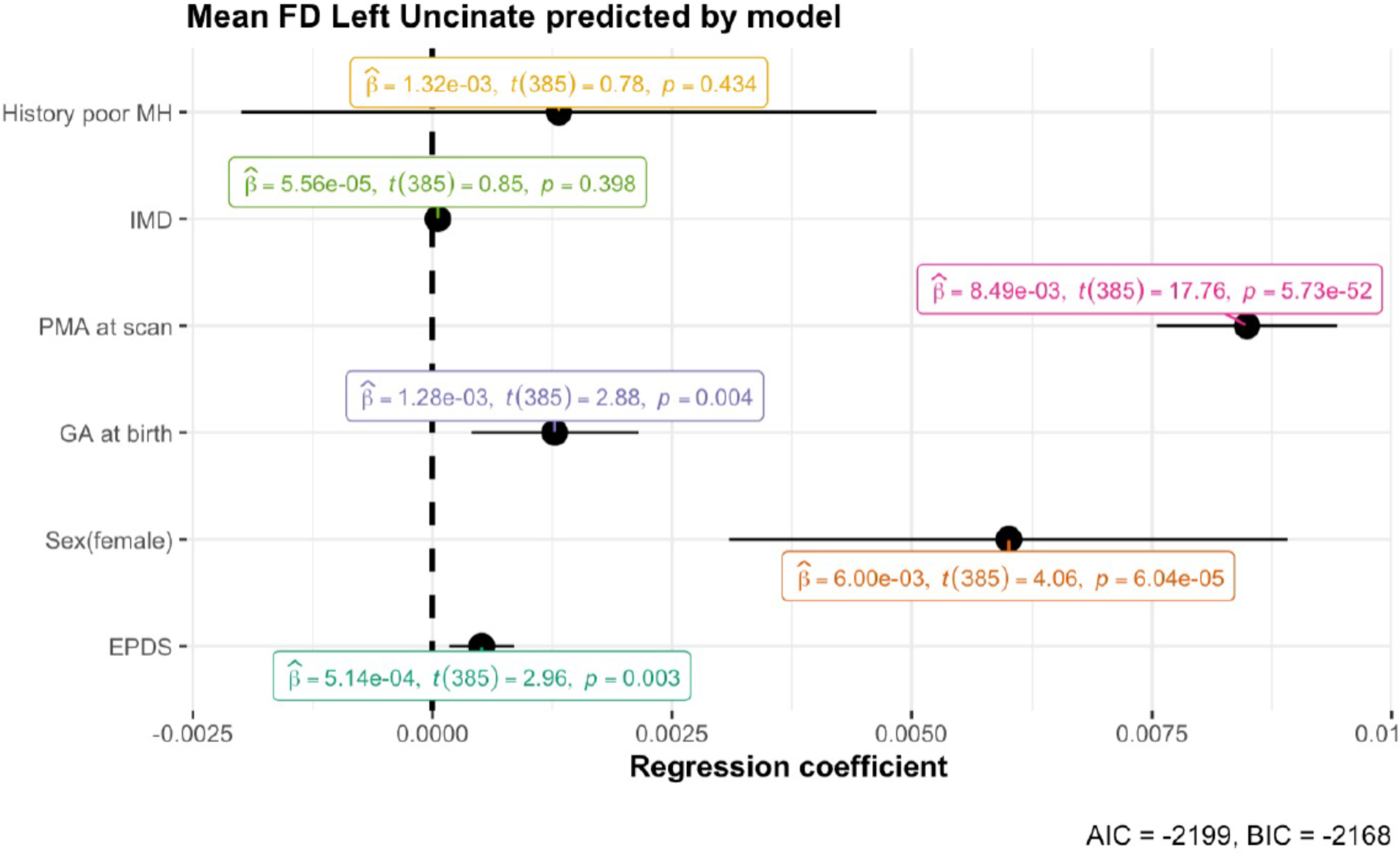
Dot-and-whisker plot for the multiple linear regression model predicting mean FD in the left uncinate fasciculus. The black dots represent the regression coefficient estimate with 95% confidence intervals. The caption contains Akaike’s Information Criterion (AIC) and the Bayesian Information Criterion (BIC), with smaller values indicating a better fit. For a model without EPDS, the AIC is -2193 and BIC is -2165, indicating a worst fit.

There was a significant EPDS by Sex interaction in left uncinate fasciculus FDC (B=-.0008, t(383)=- 2.622, p=.009), such that higher EPDS scores were associated with higher FDC in males (B=.0007, 95% CI [.0003 - .0012]) but not females (B=-.00006, 95% CI [-.0005 - .0004]). EPDS also had a significant main effect on right uncinate fasciculus FDC (B=.0004, t(384)=2.295, p=.022). These relationships did not survive correction for multiple comparisons (q=.054, q=.099).

There was no evidence for associations between EPDS and log(FC) for the uncinate fasciculus (see Supplement).

#### 3.1.2. Cingulum

EPDS scores had a positive main effect on left dorsal cingulum FDC (B =.0003, t(384)=2.098, p=.037), but this relationship did not survive correction for multiple comparisons (q=.133). There were no associations with FD or log(FDC) (see Supplement).

#### 3.1.3. Sensitivity analyses

The analysis was repeated removing n=8 participants whose mothers used SSRIs and removing n=3 women who scored >20 on the EPDS, with comparable results (see Supplement).

#### 3.1.4. Exploratory analysis on diffusion tensor imaging data

There was a positive main association between maternal EPDS and infant FA in the left (B=.0003, t(384)=2.705, p=.007) and right uncinate fasciculus (B=.0003, t(384)=2.779, p=.006), and left dorsal cingulum (B=.0003, t(384)=2.150, p=.032), and a negative association with MD in the right uncinate fasciculus (B=-.000001, t(384)=-2.402, p=.017). These did not survive correction for multiple comparisons (q values: .063, .063, .102, .144, See Supplement).

### 3.2. Infant white matter microstructure and behaviour in toddlerhood

Results from robust regression suggested that infant FD in the uncinate fasciculus had a positive main effect on Q-CHAT scores (B =105.70, p=.0007 for left, B=70.45, p=.020 for right), so that infants with higher FD in this tract had higher Q-CHAT scores at 18 months (Fig 2). The results for the left uncinate fasciculus survived FDR correction for multiple comparisons (q=.004), while results for the right uncinate fasciculus did not (q=.06). There were no FBA by sex interactions. There were no associations between infant FD in the uncinate fasciculus and CBCL internalising and externalising scores (see Supplement). These results are consistent with those obtained using traditional regression methods (see Supplement).

#### 3.2.1 Sensitivity analyses

Results from a sensitivity analysis suggested comparable results for mean FD when controlling for postnatal home environment (B=107.45, p=.0006 for left, and B=71.09, p=.020 for right uncinate fasciculus), as well as when controlling for dysfunctional parenting (B=93.34, p=.002 for left, and B=61.86, p=.036 for right uncinate fasciculus).

### 3.3. Mediation

EPDS scores predicted total Q-CHAT scores, with B =.342, t(307)=3.003, p=.003. However, there was no evidence that FD in the left uncinate fasciculus mediated the relationship between maternal EPDS and total Q-CHAT scores (Sobel test z=1.46, p=.143, bootstrapped indirect effect=.035, SE=.02, 95%CI [-.01-.08]) (see Supplement).

## 4. Discussion

We used fixel-based fibre metrics to investigate the relationship between maternal depressive symptoms and neonatal brain development. We report, for the first time, that higher severity of maternal depressive symptoms is associated with higher fibre density (FD) in the neonatal uncinate fasciculus, when controlling for infant gestational age (GA) at birth, postmenstrual age (PMA) at scan, sex, maternal socioeconomic status, and maternal history of poor mental health. These results are strengthened by an exploratory analysis using diffusion tensor imaging, which suggests the same direction of effect (i.e., increased fractional anisotropy, FA and decreased mean diffusivity, MD).

Further, higher FD in the uncinate fasciculus was also associated with social-emotional difficulties in toddlers. Results of a mediation analysis suggest that maternal depressive symptoms predict social-emotional difficulties, but there was no evidence that this relationship was mediated by uncinate fasciculus FD.

Our results are consistent with several previous dMRI studies which found increased FA and/or decreased diffusivity following exposure to maternal depression (8, 15), SSRI use (9, 78), cortisol (79) and paternal early life stress (80). Nolvi and colleagues (15) reported that increased whole-brain FA moderated the association between maternal depression and infant reactivity. However, we provide the first evidence that maternal depressive symptoms are associated with density of fibres in the uncinate fasciculus, but not fibre bundle size, in the neonatal period. In studies of rats, exposure to early stress has been shown to lead to accelerated myelination. For example, in male juvenile rats, exposure to early stress leads to accelerated myelination in the basolateral amygdala (81). It is possible maternal stress leads to accelerated axonal growth and myelination of the uncinate fasciculus, however this hypothesis requires further investigation.

Typically, increased FD, increased FA, and decreased MD during infancy suggest white matter maturation (e.g., increased axonal density) and are associated with positive outcomes (36, 68). However, our findings may provide support for the Stress Acceleration Hypothesis (82) which suggests that early adversity reprioritises developmental goals and leads to an accelerated developmental trajectory. The switch from growth to maturation (83) may offer a short-term survival advantage in a high-stress extrauterine environment (e.g., limbic areas often appear to be more mature following adversity), but may have adverse consequences in the longer term. Indeed, several structural MRI studies have reported prenatal-stress related increases in brain volumes (84–87).

Higher FA of the uncinate fasciculus in children has also been related to maternal unpredictability during infancy (88), and adolescents who had experienced increased early life trauma were more likely to be misclassified as adults based on their fronto-limbic microstructure (89). It has been proposed that accelerated maturation in response to prenatal stress aims to compensate for reduced maternal investment (90). This theory is also supported by animal studies showing that early stress is associated with precocious myelination (81) and enlarged volumes (91).

In a recent FBA study in children and adolescents (92), severity of early life stress was associated with more mature-appearing white matter structure (i.e., increased FDC), in tracts including the uncinate fasciculus. However, this was associated with fewer internalising problems in adolescence, but not late childhood (92), which the authors suggest this may be an adaptive response. More work is needed to understand longitudinal changes in brain development.

It is therefore essential to highlight the importance of the postnatal environment following exposure to prenatal stress. Early interventions have been suggested to improve outcomes for children at increased likelihood for autism (93, 94) and infants exposed to prenatal stress (95). In one such study, cognitive behavioural therapy and augmentative communication were associated with higher uncinate fasciculus mean FA in autism (96), while in a preliminary study using FBA, FC was decreased in several tracts in offspring of women who received cognitive behavioural therapy (97). Indeed, findings may be dependent on the time, duration, and intensity of the depressive symptoms. For example, it is possible that lower levels of stress are beneficial for white matter development, i.e., leading to increased FD, while higher levels of stress lead to decreased FD, in line with arguments that prenatal stress can be “both a risk and an opportunity factor” (98).

It is also possible that infants may be affected differently by prenatal stress based on pre-existing vulnerabilities. This may explain why, in a previous study conducted in extremely and very preterm babies, maternal stress was associated with higher uncinate fasciculus diffusivity (11). Future research is needed to establish whether the effects of maternal stress are different for typically developing babies and those already at risk for adverse neurodevelopment. Further, future research should examine the role of maternal health and complications during labour and delivery.

Although overall there is variability with regards to the direction of effect (99), there is evidence suggesting that white matter development in autism may be characterised by abnormal early and/or accelerated maturation followed by decreased FA later on (100–102). This is supported by our findings that increased FD in the uncinate fasciculus in infants is associated with social-emotional difficulties in toddlers. In a longitudinal study (19), young children with autism showed accelerated maturation of the uncinate fasciculus, which was associated with progression of social deficits. In a recent study in an overlapping sample (103), multi-modal cortical profiles in neonates predicted social-emotional performance at 18 months. Given the novelty of our findings, further research including longitudinal designs is required to replicate and better understand these results.

We identified a significant relationship between maternal depressive symptoms and microstructure in the neonatal uncinate fasciculus but not the cingulum bundle. Our results contrast reports that prenatal EPDS scores are associated altered cingulum microstructure in preschoolers (13) and that cingulum bundle FA in the neonatal period moderates the relationship between postpartum depression and infant reactivity at 6 months (15). However, it is important to note that Hay and colleagues assessed cingulum bundle microstructure at 4 years of age and maternal depression was measured at 3 and 6 months postpartum in Nolvi and colleagues’ study (15). These differences in age at exposure and age at assessment may account for the differing results. We have previously reported altered diffusivity in the uncinate fasciculus in infants born prematurely is associated with maternal stressful life events (11). It is therefore possible that uncinate fasciculus microstructure in the neonatal period is associated with maternal stress whereas cingulum bundle microstructure is affected at different time points.

However, this hypothesis requires further investigation with longitudinal designs.

Compared to most previous research, our study included a larger number of women with high EPDS scores (n=52). However, the EPDS only measures depressive symptoms over the previous 7 days, which means that elevated scores could have been transient in some women (104). Future research should assesses a broader range of maternal psychological symptoms, more frequently, and across the whole perinatal and early postnatal period (e.g., (105)).

Future studies are also needed to further explore the interaction between foetal sex and exposure to maternal depression. Some prior studies have reported sex differences in the white matter of offspring exposed to prenatal stress (13, 78). In our study, prenatal EPDS score was only associated with FDC in the left uncinate fasciculus in males, but this sex difference did not survive correction for multiple comparisons.

Increased maternal EPDS score during pregnancy or around the time of birth also predicted Q-CHAT scores in the first step of the mediation analysis. Children born to mothers with high EPDS scores were twice as likely to have social-emotional difficulties (22.9%) than those with low EPDS scores (11.2%) (Table 2). However, this relationship was not found to be mediated by uncinate fasciculus FD. Several factors may account for this. Firstly, it is likely that the biological mechanisms mediating the development of social-emotional traits are more complex than captured in our study design (106). Secondly, our study lacks information about postnatal mental health, so it is possible that the observed effects may be related to postnatal stress (107). Further, it is possible that maternal postnatal depressive symptoms may affect the reporting of offspring behaviour, in line with the depression-distortion hypothesis (108, 109). Future research should control for maternal postnatal mental health and ideally complement parent-reported measures of behaviour with teacher reports and observational measures.

It is unclear why in our study there were no associations with CBCL scores. One possibility is that this is related to the uncertainty of differentiating between normative behaviours and psychopathology at such an early age. However, the CBCL has good validity in infancy and toddlerhood and satisfactory predictive validity (69, 110). With regards to the Q-CHAT, reports from parents of autistic children often mention concerns around the first year of life (111, 112). However, it is important to note that the Q-CHAT has been suggested to have low predictive validity and the appearance of early social-emotional difficulties does not imply a later autism diagnosis (113), highlighting the importance of longitudinal study designs. In addition, Goh and colleagues (114) reported that higher maternal depressive symptoms were associated with a small but significant increase in maternal-reported QCHAT socio-communicative, but not behavioural, traits in healthy toddlers. Although we did not measure maternal depressive symptoms at follow-up, it is possible that mothers with persistent depressive symptoms were more likely to report socio-communicative difficulties measured with the QCHAT than behavioural difficulties on the CBCL. It is also possible that offspring are affected differently by prenatal depression based on other factors such as genetic susceptibility (115). Future studies should aim to capture individual variability in outcomes, which could be achieved using different analysis techniques such as normative modelling (116).

To our knowledge, this represents the largest study investigating the relationship between maternal depressive symptoms and neonatal white matter, as well as the first such study to use fixel-based fibre metrics, which increase microstructural and spatial specificity (36). We encourage future research to replicate these results, addressing the limitations outlined above and including even larger samples (117). Our findings have important implications for clinical practice, as a better understanding of how maternal depressive symptoms can impact offspring brain and behaviour can help inform future interventions and ensure more positive outcomes for mother and child.

## Data Availability

Imaging data and minimal demographics data are available online at http://www.developingconnectome.org/data-release/. We are unable to share further data as this would increase the risk of participant identification.

## Acknowledgements

We would like to acknowledge the contributions of our participants and their families; without whom, this work would not have been possible. We thank the nurses and midwives involved in data collection and the wider dHCP team.

This work received funding from the European Research Council under the European Union’s Seventh Framework Programme (FP7/20072013/ECR grant agreement no [319456] dHCP project) and the Medical Research Council [grant numbers MR/N013700; MR/V002465/1]. This paper represents independent research partly funded by the National Institute for Health Research (NIHR) Biomedical Research Centres at South London and Maudsley NHS Foundation Trust and at MRC Centre for Neurodevelopmental Disorders, King’s College London. This work was supported by core funding from the Wellcome/EPSRC Centre for Medical Engineering [WT203148/Z/16/Z]. DC is supported by a post-doctoral fellowship of the Flemish Research Foundation (FWO) [12ZV420N]. DB received support from a Wellcome Trust Seed Award in Science [217316/Z/19/Z]. The views expressed are those of the author(s) and not necessarily those of the MRC, the NHS, the NIHR or the Department of Health and Social Care. The funders had no involvement in the collection, analysis, or interpretation of data, in the writing of the report, or in the decision to submit the article for publication.

## Disclosures

The authors report no biomedical financial interests or potential conflicts of interest.

## Notes

### Competing Interest Statement

The authors have declared no competing interest.

### Funding Statement

This work received funding from the European Research Council under the European Union Seventh Framework Programme (FP7/20072013/ECR grant agreement no [319456] dHCP project) and the Medical Research Council [grant numbers MR/N013700 and MR/V002465/1]. This paper represents independent research partly funded by the National Institute for Health Research (NIHR) Biomedical Research Centres at South London and Maudsley NHS Foundation Trust and at MRC Centre for Neurodevelopmental Disorders at Kings College London. This work was supported by core funding from the Wellcome/EPSRC Centre for Medical Engineering [WT203148/Z/16/Z]. DC is supported by a post-doctoral fellowship of the Flemish Research Foundation (FWO) [12ZV420N]. DB received support from a Wellcome Trust Seed Award in Science [217316/Z/19/Z].

### Author Declarations

This study was approved by the UK National Research Ethics Authority (London - Riverside 14/LO/1169) and conducted in accordance with the World Medical Association Code of Ethics (Declaration of Helsinki).

