## Supplement for "Maternal depressive symptoms, neonatal white matter, and toddler social-emotional development"

*joint-first author

#### Corresponding author

Alexandra Lautarescu

Centre for the Developing Brain, Department of Perinatal Imaging and Health, School of Biomedical Engineering and Imaging Sciences, King's College London, Westminster Bridge Road, London, SE1 7EH, United Kingdom

+44 (0) 20 7188 53623

ORCID: 0000-0001-7680-8936

**Running title**: Maternal depression, infant white matter, and behaviour

The RMarkdown analysis code is available at https://osf.io/eu63s/

### Additional demographic information

##### Additional information on participant exclusion criteria

Participant exclusion criteria is provided in the manuscript, with additional information included below.

In cases where participants had more than one neonatal magnetic resonance imaging (MRI) scan, only one of the scans was included in the analysis (i.e., n=106 repeat scans were excluded). The included scan was typically the first neonatal scan, unless the first scan did not meet other inclusion criteria, in which case the second scan (n=18) or the third scan (n=1) was included instead. This was the case, for example, for preterm infants who were scanned both at birth and at term-equivalent age, when the postmenstrual age (PMA) at the first scan(s) was <37 weeks.

The Edinburgh Postnatal Depression (EPDS) questionnaire was included in the analysis if mothers completed it during pregnancy (e.g., at a visit for a fetal scan) or in the early postnatal period (i.e., at the time of the first neonatal scan), as these scores were more likely to be reflective of mood during the prenatal and early postnatal period. EPDS questionnaires completed later in the postnatal period (e.g., second or third neonatal scan) were not considered for analysis. Participants were excluded if the mothers had not completed any EPDS questionnaire during the pregnancy or at the first neonatal scan (n=100). For more information about the EPDS in this cohort, please see our previously published work (1)

Further, infants who were not singletons (e.g., twins, triplets) were excluded from further analysis (n=87).

Participants were then excluded if the diffusion MRI (dMRI) data was classed as missing (i.e., there was no dMRI data acquired) or incomplete (i.e., the number of volumes acquired was insufficient, as determined by developing Human Connectome Project, dHCP quality control measures).

N=21 participants were then excluded for failing dHCP quality control. This refers to standardised dHCP quality control procedures, which have been described elsewhere (https://biomedia.github.io/dHCP-release-notes/qc.html)

With regards to participant age, infants who were born extremely preterm (i.e., < 28 weeks gestational age, GA) or very preterm (i.e., 28-32 weeks GA) (n=46) were excluded, as well as infants who were only scanned before 37 weeks GA (n=11).

Participants were then excluded if they had major incidental findings as determined by a radiologist (e.g., major lesions, small brain < 1$^{st}$ centile) (n=22).

Visual quality control on the remaining sample led to the exclusion of 2 further participants (with areas of high signal intensity). Lastly, 1 participant was excluded due to not having a successful T2-weighted scan. The final sample consisted of n=413 participants.

EPDS scores in the participants who were excluded from further analysis were similar to the EPDS scores in the final (n=413) sample. A notable exception was the two groups of participants born prematurely (i.e., n=46 participants with gestational age at birth under 32 weeks and n=11 participants with postmenstrual age at scan under 37 weeks), where mothers scored slightly higher on the EPDS (i.e., median total EPDS score of 7).

#### Detailed information on maternal history of poor mental health

As part of the dHCP mother's questionnaire pack, participants were asked "Have you ever been treated for a mental health problem" (Yes/No). If participants answered no, no further questions were asked. If participants answered yes, the following three Yes/No questions followed: "Have you ever been under psychiatric services?", "Have you ever been admitted to hospital for a mental health problem", "Do you have a history of ADHD, bipolar disorder, depression, autism, or schizophrenia?".

To obtain a more comprehensive understanding of participant history of poor mental health, information was collated from several sources including maternal self-report, maternal electronic and hand-held notes, and records from the South London and Maudsley NHS Foundation Trust.

Based on this data, a new variable named "History of poor mental health" was created. This was coded as "Yes" if mothers disclosed having been treated for a mental health problem as part of the dHCP questionnaire, or if evidence for a history of poor mental health was found in any of the above-mentioned sources; otherwise, this was coded as "no".

Mothers with high EPDS scores were more likely to have a history of poor mental health (51.9%) compared to mothers with low EPDS scores (24.4%). The distribution of EPDS scores is presented in Fig S1.

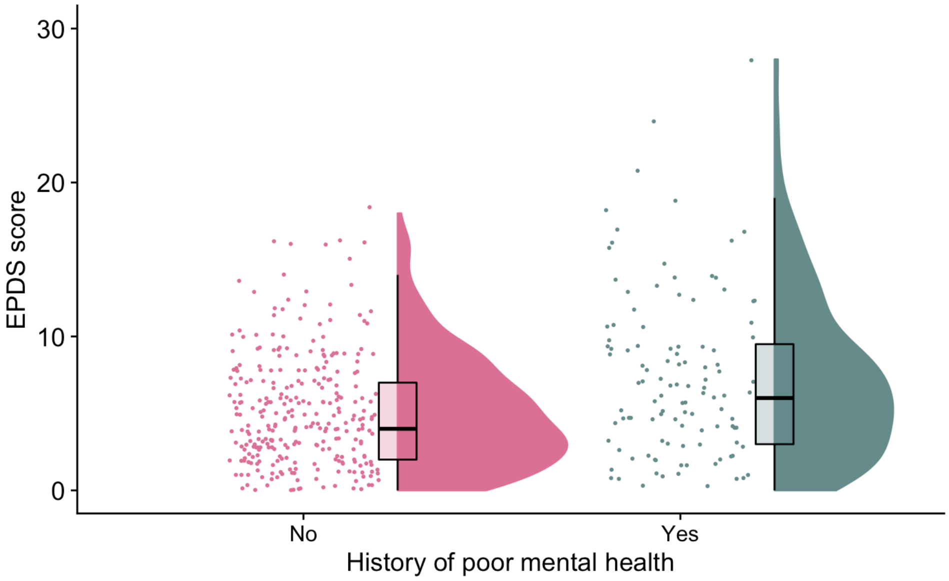


*Fig S1. Raincloud plots showing the distribution of EPDS scores for participants with and without a history of poor mental health. For each group, jittered raw data are shown on the left, boxplots with median and interquartile range are shown in the middle, and density plots are shown on the right.*

#### Additional information on participant ethnicity

In Table 1 in the main manuscript, participant ethnicity was categorized as follows:

- Asian/Asian British includes the following sub-categories: Indian, Pakistani, Bangladeshi, Chinese, Other
- Black/Black British includes the following sub-categories: African, Caribbean, Other
- Mixed ethnic group includes the following sub-categories: White and Asian, White and Black African, White and Black Caribbean, Other

### Additional methodology

#### Additional information regarding MR acquisition

Infants were monitored throughout the scan using pulse oximetry, respiratory rate, electrocardiography, and temperature. The impact of scanner noise was minimized by using earplugs moulded from a silicone-based putty placed in the external auditory meatus (President putty, Coltene Whaledent, Mahwah, NJ, USA), neonatal earmuffs (MiniMuffs, Natus Medical Inc., San Carlos, CA, USA), as well as an acoustic foam hood that was positioned over the infant to absorb noise.

#### Additional information regarding pre-processing

All T2-weighted images were motion corrected and reconstructed to a 0.8 mm isotropic resolution (2), bias field corrected (3), and brain extracted (4). Intracranial structures were segmented using Draw-EM (5,6) and total intracranial volume (ICV) was calculated by summing the volume of all tissue components, ventricles, and extracerebral CSF.

#### Additional information on segmentation for calculation of total intracranial volume

Intracranial structures were segmented into white matter, grey matter, deep grey matter, cerebrospinal fluid, ventricles, brainstem, hippocampus and amygdala and cerebellum using an extension of the Draw-EM algorithm.

### Additional information about the tracts of interest

Table S1. Regions of interest used in the delineation of the tracts

| Tract | Command | ROI (label) |
|---|---|---|
| UF-L | Include | Left middle frontal gyrus, orbital part (9) + Left inferior frontal gyrus, orbital part (15) |
| | | Left temporal pole: superior temporal gyrus (83) + Left temporal pole: middle temporal gyrus (87) |
| | Exclude | Left thalamus (77) |
| | | Left middle temporal gyrus (85) |
| | | Interhemispheric mask |
| UF-R | Include | Right middle frontal gyrus, orbital part (10) + Right inferior frontal gyrus, orbital part (16) |
| | | Right temporal pole: superior temporal gyrus (84) + Right temporal pole: middle temporal gyrus (88) |
| | Exclude | Right thalamus (78) |
| | | Right middle temporal gyrus (86) |
| | | Interhemispheric mask |
| CV-L | Include | Left parahippocampal gyrus (39) |
| | | Left posterior cingulate gyrus (35) |
| | Exclude | Interhemispheric mask |
| CV-R | Include | Right parahippocampal gyrus (40) |
| | | Right posterior cingulate gyrus (36) |
| | Exclude | Interhemispheric mask |
| CD-L | Include | Left anterior cingulate and paracingulate gyri (31) |
| | | Left posterior cingulate gyrus (35) |
| | Exclude | Interhemispheric mask |
| CD-R | Include | Right anterior cingulate and paracingulate gyri (32) |
| | | Right posterior cingulate gyrus (36) |
| | Exclude | Interhemispheric mask |

ROI=region of interest, UF-L=left uncinate fasciculus, UF-R=right uncinate fasciculus, CV-L=left ventral cingulum, CV-R=right ventral cingulum, CD-L=left dorsal cingulum, CD-R=right dorsal cingulum, Interhemispheric mask = sagittal slab 1 voxel thick through interhemispheric fissure

Note: For the uncinate fasciculus, the maximum streamline length was set at 50mm, and for the ventral and dorsal cingulum bundle, the maximum streamline length was set at 65mm.

Table S2. Fixel-based metrics for tracts of interest

| Tract | Mean (SD) FD | Mean (SD) log(FC) | Mean (SD) FDC |
|---|---|---|---|
| Left uncinate fasciculus | .21 (.02) | -.02 (.08) | .20 (.03) |
| Right uncinate fasciculus | .20 (.02) | -.02 (.08) | .20 (.03) |
| Left ventral cingulum | .24 (.02) | -.04 (.08) | .23 (.02) |
| Right ventral cingulum | .24 (.02) | -.03 (.08) | .23 (.02) |
| Left dorsal cingulum | .22 (.02) | -.03 (.09) | .21 (.02) |
| Right dorsal cingulum | .20 (.02) | -.04 (.09) | .20 (02) |

SD=standard deviation, FD=fibre density, FC=fibre cross-section, FDC=fibre density and cross-section

Table S3. Diffusion tensor imaging metrics for tracts of interest

| Tract | Mean (SD) FA | Mean (SD) MD |
|---|---|---|
| Left uncinate fasciculus | .15 (.01) | .0013 (.00006) |
| Right uncinate fasciculus | .15 (.01) | .0013 (.00007) |
| Left ventral cingulum | .18 (.01) | .0013 (.00005) |
| Right ventral cingulum | .18 (.01) | .0013 (.00005) |
| Left dorsal cingulum | .18 (.01) | .0012 (.00005) |
| Right dorsal cingulum | .18 (.02) | .0012 (.00005) |

SD=standard deviation, FA=fractional anisotropy, MD=mean diffusivity

Table S4. Spearman's rank correlation matrix for the associations between fixel-based metrics and diffusion tensor imaging measures. Values represent correlation coefficients for all correlations under p<.05;

| | | UF-L FA | UF-L MD | UF-R FA | UF-R MD | CV-L FA | CV-L MD | CV-R FA | CV-R MD | CD-L FA | CD-L MD | CD-R FA | CD-R MD |
|---|---|---|---|---|---|---|---|---|---|---|---|---|---|
| UF-L | FD | rho=.96** | rho=-.92** | | | | | | | | | | |
| | FDC | rho=.77** | rho=-.62** | | | | | | | | | | |
| | Log(FC) | rho=-.007 p=.894 | rho=.18** | | | | | | | | | | |
| UF-R | FD | | | rho=.97** | rho=-.93** | | | | | | | | |
| | FDC | | | rho=.77** | rho=-.63** | | | | | | | | |
| | Log(FC) | | | rho=-.06 rho=.261 | rho=.21** | | | | | | | | |
| CV-L | FD | | | | | rho=.90** | rho=-.89** | | | | | | |
| | FDC | | | | | rho=.67** | rho=-.52** | | | | | | |
| | Log(FC) | | | | | rho=.07 p=.173 | rho=.07 p=.165 | | | | | | |
| CV-R | FD | | | | | | | rho=.92** | rho=-.90** | | | | |
| | FDC | | | | | | | rho=.68** | rho=-.56** | | | | |
| | Log(FC) | | | | | | | rho=.06 p=.246 | rho=.05 p=.354 | | | | |
| CD-L | FD | | | | | | | | | rho=.90** | rho=-.87** | | |
| | FDC | | | | | | | | | rho=.58** | rho=-.50** | | |
| | Log(FC) | | | | | | | | | rho=-.05 p=.279 | rho=.10 p=.040 | | |
| CD-R | FD | | | | | | | | | | | rho=.90** | rho=-.88** |
| | FDC | | | | | | | | | | | rho=.58** | rho=-.53** |
| | Log(FC) | | | | | | | | | | | rho=-.06 p=.240 | rho=.10 p=.038 |

UF-L=left uncinate fasciculus, UF-R=right uncinate fasciculus, CV-L=left ventral cingulum, CV-R=right ventral cingulum, CD-L=left dorsal cingulum, CD-R=right dorsal cingulum, FA=fractional anisotropy, MD=mean diffusivity, FD=fibre density, FC=fibre cross-section, FDC=fibre density and cross-section.

Note: ** denotes p<.001; otherwise the p value is provided in table.

As a quality control measure, the relationship between fixel-based metrics and PMA at scan was examined. For all tracts, PMA at scan was positively associated with mean fibre density, FD (rho between .53 and .74, p<.001), mean fibre density and cross-section, FDC (rho between .70 and .80, p<.001) and the log of fibre-cross section, log(FC) (rho between .28 and .45, p<.001). An example of this association is provided in Figure S2.

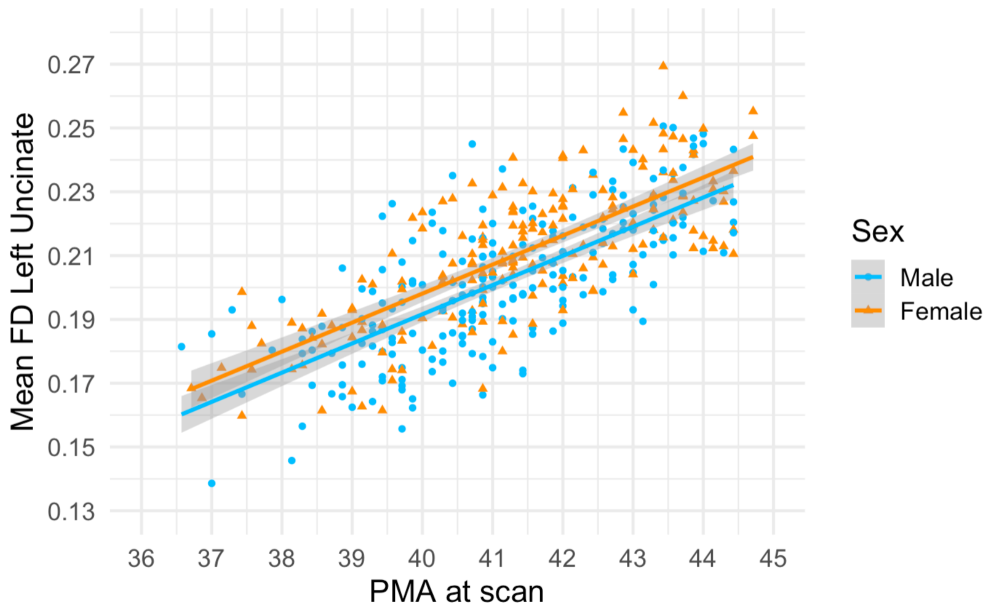


*Fig S2. Association between mean fibre density (FD) in the left uncinate fasciculus and infant postmenstrual age (PMA) at scan*

#### Associations between potential covariates and variables of interest

In the interest of transparency, the decision-making process for including covariates in the regression models is summarised below.

##### Potential covariates for the "EPDS – FBA metrics" models

Potential covariates were chosen based on previous literature, and their relationship with variables of interest was assessed using a Spearman's rank correlation matrix.

The variables that were included in all models are as follows: GA at birth, PMA at scan, infant sex, maternal index of multiple deprivation (IMD) at enrolment, maternal history of mental health. In addition, for FDC and log(FC) models, ICV was also included as a covariate. The reasoning behind including these variables is detailed below:

- GA at birth was included in the models as it was positively associated with mean FD and FDC in the tracts of interest (rho values between .35 and .51; all p<.001); it was not associated with log(FC) or with EPDS scores.

- PMA at scan was included in the models as it was positively associated with EPDS scores (rho=.12, p=.015) and mean FD, FDC and log(FC) in all tracts of interest (rho values between .28 and .80, all p<.001).
- Infant sex was included in the models based on previous literature suggesting that maternal depression may impact male and female offspring differently (7–9), as well as established knowledge that male brains are larger than female brains, which can confound differences in brain anatomy (10,11).
- Maternal IMD score at study enrolment was negatively associated with log(FC) in the left ventral cingulum (rho=-.14, p=.005), right ventral cingulum (rho= -.15, p=.002), left dorsal cingulum (rho=-.12, p=.020) and right dorsal cingulum (rho=-.10, p=.043). There was no association with maternal EPDS score, or with other fixel-based fibre metrics. This variable was included in the models based on this, as well as previously established associations with maternal mental health and child development (12–14)
- Maternal history of poor mental health was included in the models, as it was associated with EPDS scores (W=13235, p<.001), with women with a history of poor mental health having higher EPDS scores (mean=7.24, SD=5.40) than those without a history (mean=5.08, SD=3.69).
- In addition, intracranial volume (ICV) was included in models for FDC and log(FC) based on literature suggesting that ICV is positively associated with these metrics across white matter regions (15). FD is a measure of density and should be unaffected by macroscopic morphology. In our sample, ICV was associated with FDC and log(FC) in all tracts of interest (p values between <.001 and .02).

Several variables were considered but not included in the models, as below:

- Maternal age was not associated with any of the variables of interest and was not included in the models.
- Pre-pregnancy maternal body mass index (BMI) at enrolment was not included in the models as it was not associated with any of the variables of interest, with the exception of a weak association with mean log(FC) in the left uncinate fasciculus (rho=-.10, p=.042)
- Infant birth weight was associated with all tracts of interest (rho values between .16 and .39, p<.001) but it was strongly associated with GA at birth, so it was not included in the models to avoid issues related to multicollinearity.

"The relationship between variables of interest and other potential variables was assessed post-hoc, but these were not included in sensitivity analyses, as below.

- For babies with suspected intrauterine growth restriction (IUGR) (n=19), maternal EPDS scores were similar (mean=5.05, SD=3.85) to those without (mean=5.7, SD=4.38). Values were also similar between these two groups for mean FD in the left uncinate fasciculus (both mean=0.2, SD=.02) and right uncinate fasciculus (both mean=0.2, SD=.02).
- For cases where the mother had past medical health problems (including asthma, diabetes, Crohn disease, epilepsy, chronic hypertension, thrombocytopenia) (n=233), maternal EPDS scores were similar (mean=5.62, SD=4.28) to those without past medical health problems (mean=5.76, SD=4.43). Values were also similar between these two groups for mean FD in the left uncinate fasciculus (both mean=0.2, SD=.02) and right uncinate fasciculus (both mean=0.2, SD=.02).
- Maternal parity in this cohort had a median of 0, ranging between 0 and 6. These values were the same when splitting the cohort into high EPDS and low EPDS. Results of a Spearman's rank correlation suggested that maternal EPD and maternal parity are not significantly correlated (rho=-.08, p=.085).

#### Potential covariates for the "Fixel based metrics – behaviour" models

Potential covariates were chosen based on previous literature, and their relationship with variables of interest was assessed using a Spearman's rank correlation matrix.

The variables that were included in all models are as follows: infant GA at birth, infant PMA at scan, infant sex, maternal IMD at 18-month assessment, child corrected age at assessment in months, Bayley's cognitive composite score, and maternal age. The reasoning behind including these variables is detailed below:

- GA at birth was included in the models as it was associated with the fixel based metrics: mean FD in the left uncinate fasciculus (rho=.51, p<.001) and right uncinate fasciculus (rho=.50, p<.001).
- PMA at scan was included in the models as it was associated with the fixel based metrics: mean FD in the left uncinate fasciculus (rho=.76, p<.001) and right uncinate fasciculus (rho=.75, p<.001). Given the high correlation between PMA and FD, variance inflation factors (VIF) were assessed prior to including both as predictors in regression models, to ensure that multicollinearity is not an issue. PMA at scan was also associated with CBCL internalising t-score (rho=-.13, p=.020) and Q-CHAT total score (rho=-.21, p<.001).
- Infant sex was included in the models; the reasoning is as detailed in the previous section

- Maternal IMD score at 18 month was included in the models based on previous literature as detailed in the previous section. In the current sample, it was associated with total Q-CHAT score (rho=.16, p=.005)
- Child corrected age at assessment was not associated with any of the variables of interest, but it was included in the model to control for age-related potential differences in behavioural scores.
- Bayley's cognitive composite score was included in the models; it was associated with Q-CHAT scores (rho=-.29, p<.001), CBCL internalising scores (rho=-.20, p<.001), mean FD for the left uncinate fasciculus (rho=.14, p=.015), and mean FD for the right uncinate fasciculus (rho=.13, p=.026)
- Maternal age was included in the models as it was associated with CBCL internalising score (rho=-.15, p=.007).

Maternal IMD score at the neonatal timepoint was not included in the models, to avoid issues related to multicollinearity, as it was strongly associated with maternal IMD score at 18 months (rho=.78, p<.001).

#### Multiple regression: Maternal depression and infant white matter microstructure

##### Assumptions for multiple regression

The GVLMA package in R was used to assess linear model assumptions, with the output being represented by a single global test, as well as specific directional tests for assessing skewness, kurtosis, nonlinear link function and heteroscedasticity. If the GVLMA results suggested that some assumptions were not met, these were investigated in more detail using specific tests and residual plots as detailed below.

###### Assumptions for FD models

Results from the GVLMA package suggested that all assumptions for multiple regression were met for all FD models, with the exception of the right ventral cingulum FD, so further tests were conducted on this model.

The GVLMA results for the right ventral cingulum FD model suggested that the assumption of homoscedasticity was not met. The plot of residuals vs fitted values was checked, and it did not suggest heteroscedasticity. The plot for scale-location was a relatively straight, horizontal line, also suggesting homoscedasticity. To confirm this, two formal tests were conducted; the non-constant variance (NCV) score test was not significant (Chi-square=.245, p=.620) and neither was the Breusch-Pagan test (BP=12.44, p=.052). Overall, these suggested that the assumption of homoscedasticity was met in this model.

#### Assumptions for FDC models

Results from the GVLMA package suggested that all assumptions for multiple regression were met for all FDC models.

#### Assumptions for Log(FC) models

For the Log(FC) models, GVLMA results suggested that assumptions for multiple regression were not met, and these are examined in more detail below.

For all models, there were no issues with multicollinearity (largest variance inflation factor, VIF was for the ICV variable) and no anomalous data (no Cook's distance value above 1). The outlier tests suggested potential outliers, but visual inspection of added-variable plots did not suggest that removing any outliers would substantially affect the slopes. Additional information for each of the log(FC) models is presented in Table S5.

Overall, log(FC) models showed slight deviations from linearity (driven by the ICV variable).

Table S5. Summary of assumptions check for log(FC) models

| Model | Linearity | Homoscedasticity | Normality |
|---|---|---|---|
| UF-L | *Slight deviations from linearity*<br>Visual inspection of plots suggested no major deviations<br>Tukey's test was significant (p<.001), driven by ICV variable (p=.003). | *Assumption met*<br>NCV not significant (p=.640)<br>BP significant (BP=17.92, p=.012).<br>Visual inspection of fitted vs residuals plot - ok<br>Visual inspection of scale-location plot - ok | *Assumption met.*<br>Residuals –normally distributed<br>Median residuals = .001<br>Q-Q plot – straight line<br>Shapiro-Wilk not significant (W=.995, p=.290). |
| UF-R | *Slight deviations from linearity*<br>Visual inspection of plots suggested no major deviations.<br>Tukey's test was significant (p=.042), driven by ICV variable (p=.029). | *Assumption met*<br>NCV not significant (p=.724)<br>BP not significant (BP=10.11, p=.182)<br>Visual inspection of fitted vs residuals plot - ok<br>Visual inspection of scale-location plot - ok | *Assumption met.*<br>Residuals –normally distributed<br>Median residuals = .002<br>Q-Q plot – straight line<br>Shapiro-Wilk significant (W=.992, p=.034)<br>Large sample size – analysis should be robust to violations of normality |
| CD-L | *Slight deviations from linearity*<br>Visual inspection of plots suggested no major deviations.<br>Tukey's test was significant (p<.001), driven by ICV variable (p=.002). | *Assumption met*<br>NCV not significant (p=.141)<br>BP not significant (BP=8.32, p=.305)<br>Visual inspection of fitted vs residuals plot - ok<br>Visual inspection of scale-location plot - ok | *Assumption met.*<br>Residuals –normally distributed<br>Median residuals = .002<br>Q-Q plot – straight line<br>Shapiro-Wilk not significant (W=.993, p=.092) |
| CD-R | *Slight deviations from linearity*<br>Visual inspection of plots suggested no major deviations.<br>Tukey's test was significant (p<.001), driven by ICV variable (p<.001). | *Assumption met*<br>NCV not significant (p=.206)<br>BP not significant (BP=5.23, p=.631)<br>Visual inspection of fitted vs residuals plot - ok<br>Visual inspection of scale-location plot - ok | *Assumption met.*<br>Residuals –normally distributed<br>Median residuals =-.0006<br>Q-Q plot – straight line<br>Shapiro-Wilk not significant (W=.996, p=.479) |
| CV-L | *Slight deviations from linearity*<br>Visual inspection of plots suggested no major deviations.<br>Tukey's test was significant (p<.001), driven by ICV variable (p=.007) | *Assumption met*<br>NCV not significant (p=.939)<br>BP not significant (BP=2.30, p=.941)<br>Visual inspection of fitted vs residuals plot - ok<br>Visual inspection of scale-location plot - ok | *Assumption met.*<br>Residuals –normally distributed<br>Median residuals =.0002<br>Q-Q plot – straight line<br>Shapiro-Wilk not significant (W=.996, p=.422) |
| CV-R | *Slight deviations from linearity*<br>Visual inspection of plots suggested no major deviations.<br>Tukey's test was significant (p=.001), driven by ICV variable (p=.003) | *Assumption met*<br>NCV not significant (p=.333)<br>BP not significant (BP=4.12, p=.766)<br>Visual inspection of fitted vs residuals plot - ok<br>Visual inspection of scale-location plot - ok | *Assumption met.*<br>Residuals –normally distributed<br>Median residuals =-.0009<br>Q-Q plot – straight line<br>Shapiro-Wilk not significant (W=.995, p=.268) |

UF-L = left uncinate fasciculus, UF-R=right uncinate fasciculus, CD-L=left dorsal cingulum, CD-R=right dorsal cingulum, CV-L=left ventral cingulum, CV-R=right ventral cingulum, ICV=intracranial volume, NCV=non-constant variance test, BP=Breusch-Pagan

##### Additional analysis excluding the ICV covariate (for log(FC) models)

As some of the assumptions for log(FC) models were not fully met (i.e., slight deviations from linearity), the analyses were repeated excluding the ICV variable from the models. These new models met all assumptions for multiple regression, and results were similar to those in the main analysis, with no relationship between EPDS scores and mean log(FC) for the left uncinate fasciculus (B=-.0007, p=.380), right uncinate fasciculus (B=-.0007, p=.377), left dorsal cingulum (B=.0002, p=.808), right dorsal cingulum (B=0002, p=.799), left ventral cingulum (B=-.0002, p=.807), and right ventral cingulum (B=-.0001, p=.873).

#### Sensitivity analyses

##### Sensitivity analysis: Excluding n=8 participants with SSRI exposure

N=8 women were taking SSRI medication during their pregnancy (n=5 sertraline, n=2 fluoxetine, n=1 citalopram, n=1 unspecified). The median EPDS scores for these women was 8.5 (range 0-19). The regression models were repeated excluding these participants from the analysis, and the results were comparable (Table S6).

Table S6 Regression models excluding n=8 participants with SSRI exposure

| Tract | Metric | EPDS model | | | EPDS x Sex (female) model | | | Model fit | |
|---|---|---|---|---|---|---|---|---|---|
| | | B | t | p (q) | B | t | p (q) | $R^2$ | $F_{(df)}$ |
| UF-L | FD | .0005 | 2.98 | .003 (.027)* | | | | .58 | $F_{(6,379)}$=89 |
| | FDC | | | | -.0008 | -2.36 | .019 (.112) | .73 | $F_{(8,377)}$=132.9 |
| | Log(FC) | -.0007 | -1.40 | .163 | | | | .72 | $F_{(7,378)}$=140.7 |
| UF-R | FD | .0006 | 2.96 | .003 (.027)* | | | | .57 | $F_{(6,379)}$=87.04 |
| | FDC | .0004 | 2.24 | .025 (.112) | | | | .74 | $F_{(7,378)}$=158.8 |
| | Log(FC) | -.0007 | -1.44 | .152 | | | | .74 | $F_{(7,378)}$=154.3 |
| CD-L | FD | .0003 | 1.44 | .150 | | | | .34 | $F_{(6,379)}$=34.54 |
| | FDC | .0003 | 2.02 | .044 (.158) | | | | .65 | $F_{(7,378)}$=102 |
| | Log(FC) | .0002 | 0.47 | .639 | | | | .78 | $F_{(7,378)}$=199 |
| CD-R | FD | .0002 | 1.14 | .254 | | | | .37 | $F_{(6,379)}$=38.25 |
| | FDC | .0003 | 1.76 | .079 | | | | .66 | $F_{(7,378)}$=109.6 |
| | Log(FC) | .0004 | 0.76 | .447 | | | | .74 | $F_{(7,378)}$=155 |
| CV-L | FD | .0002 | 1.21 | .228 | | | | .40 | $F_{(6,379)}$=44.48 |
| | FDC | .0002 | 1.32 | .188 | | | | .67 | $F_{(7,378)}$=110.3 |
| | Log(FC) | -.00004 | -.09 | .929 | | | | .80 | $F_{(7,378)}$=222.1 |
| CV-R | FD | .0002 | 1.13 | .258 | | | | .44 | $F_{(6,379)}$=51.11 |
| | FDC | .0002 | 1.16 | .247 | | | | .68 | $F_{(7,378)}$=119.4 |
| | Log(FC) | .00006 | 0.14 | .892 | | | | .79 | $F_{(7,378)}$=210.3 |

SSRI=selective serotonin reuptake inhibitors, UF-L=left uncinate fasciculus, UF-R=right uncinate fasciculus, CD-L=left dorsal cingulum, CD-R=right dorsal cingulum, CV-L=left ventral cingulum, CV-R=right ventral cingulum, FD=fibre density, FC=fibre cross-section, FDC=fibre density and cross-section

Note: All R squared models are adjusted and significant at p<.001. * denotes q values that are significant after FDR correction for multiple comparisons.

#### Sensitivity analysis: Excluding n=3 women who scored > 20 on the EPDS

N=3 women scored very high on the EPDS (i.e. 21, 24, and 28). The analysis was repeated excluding these women, and the results were comparable (Table S7) (Fig S3).

Table S7. Regression models excluding n=3 participants with high EPDS scores

| Tract | Metric | EPDS model | | | EPDS x Sex (female) model | | | Model fit | |
|---|---|---|---|---|---|---|---|---|---|
| | | **B** | **t** | **p (q)** | **B** | **t** | **p (q)** | $R^2$ | $F_{(df)}$ |
| UF-L | FD | .0006 | 2.98 | .003 (.027)* | | | | .57 | $F_{(6,382)}$=87.5 |
| | FDC | | | | -.0009 | -2.60 | .01 (.060) | .73 | $F_{(8,380)}$=131.2 |
| | Log(FC) | | | | -.002 | -2.00 | .046 (.165) | .72 | $F_{(8,380)}$=124.7 |
| UF-R | FD | .0006 | 2.99 | .003 (.027)* | | | | .57 | $F_{(6,382)}$=85.37 |
| | FDC | .0004 | 2.27 | .024 (.108) | | | | .74 | $F_{(7,381)}$=154.7 |
| | Log(FC) | -.0008 | -1.43 | .155 | | | | .74 | $F_{(7,381)}$=155.1 |
| CD-L | FD | .0003 | 1.41 | .160 | | | | .33 | $F_{(6,382)}$=32.99 |
| | FDC | .0003 | 1.83 | .067 | | | | .64 | $F_{(7,381)}$=99.07 |
| | Log(FC) | .0003 | 0.51 | .609 | | | | .78 | $F_{(7,381)}$=197.4 |
| CD-R | FD | .0003 | 1.53 | .127 | | | | .36 | $F_{(6,382)}$=37.47 |
| | FDC | .0003 | 1.69 | .092 | | | | .66 | $F_{(7,381)}$=106.7 |
| | Log(FC) | .0002 | 0.35 | .729 | | | | .73 | $F_{(7,381)}$=154.6 |
| CV-L | FD | .0002 | 0.99 | .325 | | | | .39 | $F_{(6,382)}$=43.2 |
| | FDC | .0001 | 0.79 | .430 | | | | .66 | $F_{(7,381)}$=106.4 |
| | Log(FC) | -.0002 | -.33 | .745 | | | | .80 | $F_{(7,381)}$=219.4 |
| CV-R | FD | .0002 | 1.15 | .252 | | | | .43 | $F_{(6,382)}$=49.91 |
| | FDC | .0001 | 0.72 | .472 | | | | .68 | $F_{(7,381)}$=116.2 |
| | Log(FC) | -.00007 | -0.16 | .875 | | | | .79 | $F_{(7,381)}$=210.4 |

UF-L=left uncinate fasciculus, UF-R=right uncinate fasciculus, CD-L=left dorsal cingulum, CD-R=right dorsal cingulum, CV-L=left ventral cingulum, CV-R=right ventral cingulum, FD=fibre density, FC=fibre cross-section, FDC=fibre density and cross-section

Note: All R squared models are adjusted and significant at p<.001. * denotes q values that are significant after FDR correction for multiple comparisons.

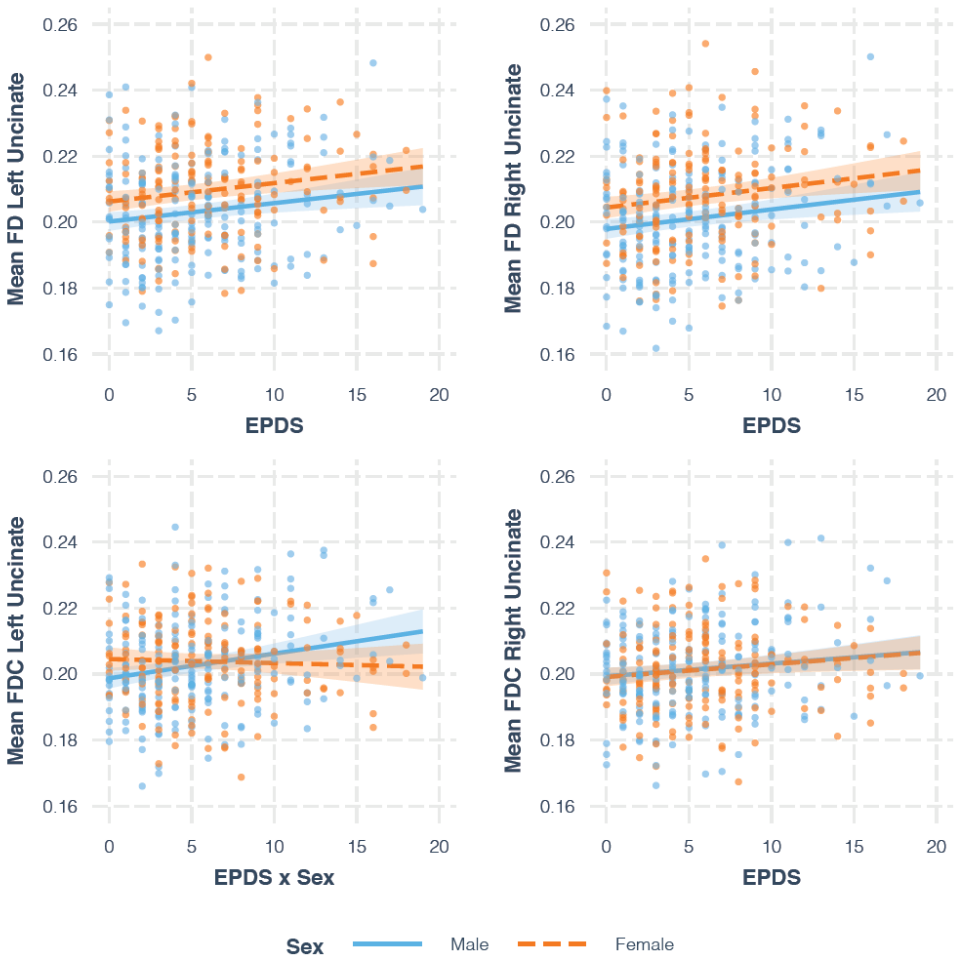


*Fig S3. Plots showing the relationship between EPDS and uncinate fasciculus FD and FDC, controlling for effect of covariates, in a sample excluding EPDS scores > 20. For mean FD left and right uncinate fasciculus and mean FDC right uncinate fasciculus, the main model is plotted. For mean FDC left uncinate fasciculus, the model with EPDS x Sex interaction is plotted.*

#### Exploratory analysis on diffusion tensor imaging data

Given that research into the effects of maternal stress on white matter microstructure has predominantly been done based on the diffusion tensor model, an exploratory analysis was conducted to investigate the relationship between maternal depression and FA and MD in the tracts of interest.

EPDS scores had a positive main effect on left uncinate fasciculus FA (B=.0003, p=.007), right uncinate fasciculus FA (B=.0003, p=.006) and left dorsal cingulum FA (B=.0003, p=.032), so that infants of mothers with higher EPDS scores tended to have higher FA. EPDS scores also had a negative main effect on MD for the right uncinate fasciculus (B=-.000001, p=.017). None of these relationships survived correction for multiple comparisons (See Table S8 and Fig S4)

Table S8. Associations between maternal EPDS and infant diffusion tensor imaging metrics

| | | EPDS model | | | EPDS x Sex (female) model | | | Model fit | |
|---|---|---|---|---|---|---|---|---|---|
| **Tract** | **Metric** | **B** | **t** | **p (q)** | **B** | **t** | **p (q)** | **$R^2$** | **$F_{(df)}$** |
| UF-L | FA | .0003 | 2.71 | .007 (.063) | | | | .55 | $F_{(7,384)}$=68.79 |
| | MD | < -.0001 | -1.82 | .070 | | | | .56 | $F_{(7,384)}$=70.9 |
| UF-R | FA | .0003 | 2.78 | .006 (.063) | | | | .58 | $F_{(7,384)}$=76.9 |
| | MD | < -.0001 | -2.40 | .017 (.102) | | | | .55 | $F_{(7,384)}$=69.14 |
| CD-L | FA | .0003 | 2.15 | .032 (.144) | | | | .29 | $F_{(7,384)}$=23.65 |
| | MD | < -.0001 | -1.36 | .175 | | | | .37 | $F_{(7,384)}$=34.42 |
| CD-R | FA | .0002 | 1.59 | .112 | | | | .30 | $F_{(7,384)}$=25.04 |
| | MD | < -.0001 | -1.66 | .098 | | | | .38 | $F_{(7,384)}$=35.82 |
| CV-L | FA | .0002 | 1.81 | .071 | | | | .33 | $F_{(7,384)}$=28.2 |
| | MD | < -.0001 | -1.42 | .157 | | | | .41 | $F_{(7,384)}$=40.17 |
| CV-R | FA | .0002 | 1.35 | .178 | | | | .34 | $F_{(7,384)}$=29.99 |
| | MD | < -.0001 | -1.45 | .149 | | | | .45 | $F_{(7,384)}$=47.57 |

UF-L=left uncinate fasciculus, UF-R=right uncinate fasciculus, CD-L=left dorsal cingulum, CD-R=right dorsal cingulum, CV-L=left ventral cingulum, CV-R=right ventral cingulum, FA=fractional anisotropy, MD=mean diffusivity,

Note: All R squared models are adjusted and significant at p<.001. * denotes q values that are significant after FDR correction for multiple comparisons.

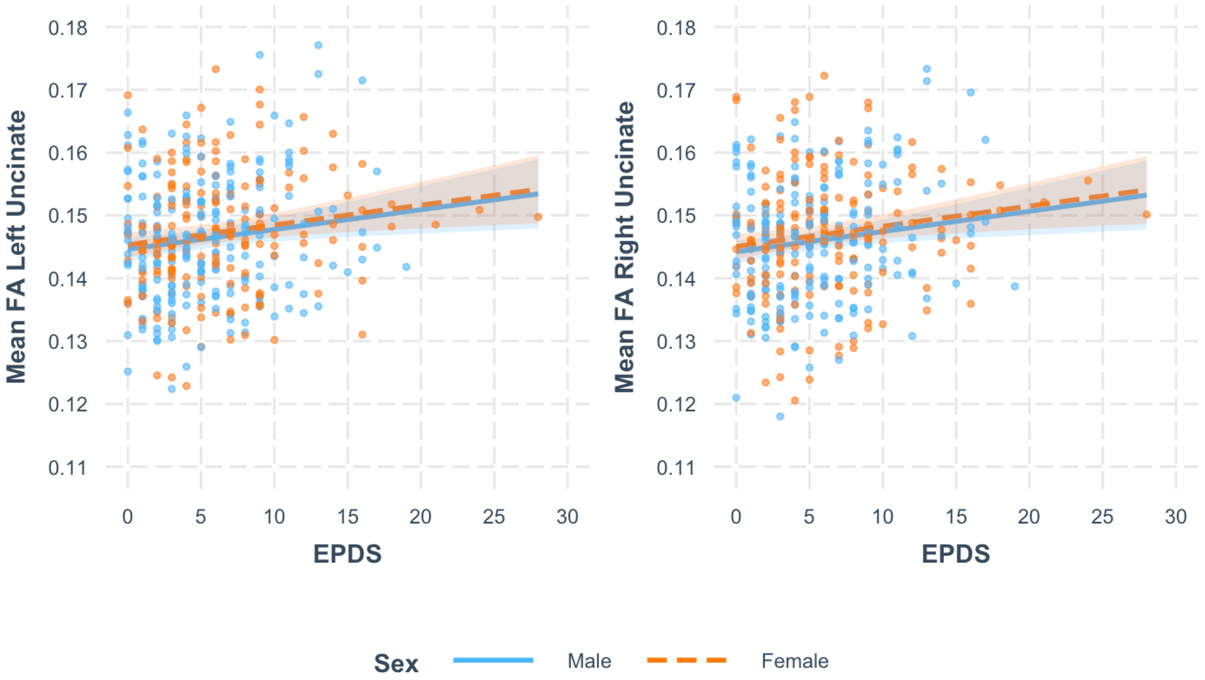


Fig S4. *Plots showing the relationship between EPDS and FA in the left and right uncinate fasciculus, controlling for effect of covariates.*

### Multiple regression: Infant white matter microstructure and toddler behaviour

#### Summary of results using traditional regression methods

Assumptions for multiple regression were not fully met (as evidenced below), so the main analysis was conducted using robust linear regression (lmrob function from the robustbase package in R). To enable comparison, results from an analysis using traditional multiple regression are reported below.

The analyses conducted with traditional regression methods and robust regression methods revealed similar results, with methods suggesting an association between Q-CHAT and FD in the left uncinate fasciculus. There was no FD by Sex interaction. CBCL internalizing and externalizing behaviour was not accurately predicted by the models, suggesting that variables other than those included (i.e. infant sex, GA at birth, PMA at scan, maternal IMD, corrected age at assessment, cognitive composite score, maternal age) better explain the variance in toddler behaviour at this age. Further research is required to understand this. The results from the robust regression and traditional linear regression are briefly presented in Tables S9 and S10.

Table S9. Robust regression: Associations between infant white matter and toddler behaviour

| Model | FD models | | | FD x Sex (female) model | | | Model fit |
|---|---|---|---|---|---|---|---|
| | **B** | **t** | **p (q)** | **B** | **t** | **p (q)** | **$R^2$** |
| Q-CHAT FD UF-L | 105.70 | 3.43 | .0007 (.004)* | | | | .17 |
| Q-CHAT FD UF_R | 70.45 | 2.33 | .020 (.06) | | | | .16 |
| CBCL Int FD UF-L | 63.23 | 1.52 | .130 | | | | .09 |
| CBCL Int FD UF-R | 59.45 | 1.46 | .144 | | | | .09 |
| CBCL Ext FD UF-L | 66.61 | 1.60 | .110 | | | | .01 |
| CBCL Ext FD UF-R | 54.74 | 1.41 | .159 | | | | .01 |

UF-L=left uncinate fasciculus, UF-R=right uncinate fasciculus, Int=Internalising, Ext=Externalising, FD=fibre density

Note: All R squared models are adjusted and significant at p<.001. * denotes q values that are significant after FDR correction for multiple comparisons.

Table S10. Traditional linear regression: Associations between FD and toddler behaviour

| Model | FD models | | | FD x Sex (female) model | | | Model fit | |
|---|---|---|---|---|---|---|---|---|
| | **B** | **t** | **p (q)** | **B** | **t** | **p (q)** | **$R^2$** | **$F_{(df)}$** |
| Q-CHAT FD UF-L | 112.05 | 3.51 | .0005 (.003)* | | | | .19 | $F_{(8,283)}$=9.50 |
| Q-CHAT FD UF_R | 74.35 | 2.42 | .016 (.048)* | | | | .17 | $F_{(8,283)}$=8.52 |
| CBCL Int FD UF-L | 67.63 | 1.75 | .081 | | | | .09 | $F_{(8,283)}$=4.70 |
| CBCL Int FD UF-R | 60.77 | 0.07 | .099 | | | | .09 | $F_{(8,283)}$=4.65 |
| CBCL Ext FD UF-L | 56.63 | 1.50 | .136 | | | | .006 | $F_{(8,283)}$=1.22 |
| CBCL Ext FD UF-R | 46.08 | 1.28 | .203 | | | | .004 | $F_{(8,283)}$=1.14 |

UF-L=left uncinate fasciculus, UF-R=right uncinate fasciculus, Int=Internalising, Ext=Externalising, FD=fibre density

Note: All R squared models are adjusted and significant at p<.001. * denotes q values that are significant after FDR correction for multiple comparisons

#### Assumptions for regression using traditional regression methods

The GVLMA package in R was used to assess linear model assumptions, with the output being represented by a single global test, as well as specific directional tests for assessing skewness, kurtosis, nonlinear link function and heteroscedasticity. If the GVLMA results suggested that some assumptions were not met, these were investigated in more detail using specific tests and residual plots as detailed below.

For all models, there were no issues with multicollinearity (largest VIF was for the PMA at scan variable). There were a few potential influential outliers, but no Cook's value above 1. For the Q-CHAT models, the added variable plot did not suggest any observations that, when removed, would significantly change the slope for the relationships between variables. For the CBCL internalising models, plots suggested 1 potential influential outlier for PMA at scan, and several potential influential outliers for maternal age. A summary of the assumptions check for the Q-CHAT and CBCL internalising models is presented in Table S11. For the CBCL externalising models, all assumptions for multiple regression were met, but models accounted only for a very small percentage of variance in scores, suggesting that other variables than the ones included in the model would better predict scores on this measure.

As the assumption of homoscedasticity was not met for the Q-CHAT models, there was a chance that standard error estimates associated with regression coefficients are no longer reliable, affecting the t and p values. A heteroscedasticity corrected covariance matrix was calculated, and results supported the associations between mean FD in the left uncinate fasciculus and total Q-CHAT score (B=112.05, p=.0003) and mean FD in the right uncinate fasciculus and total Q-CHAT score (B=74.35, p=.013, not surviving correction for multiple comparisons).

Table S11. Summary of assumptions check for Q-CHAT and CBCL internalising models

| Model | Linearity | Homoscedasticity | Normality |
|---|---|---|---|
| Q-CHAT | | | |
| FD UF-L | *Slight deviations from linearity*<br>Visual inspection of plots suggested no major deviations<br>Tukey's test was significant (p<.001), driven by the Bayley's cognitive score (p<.001) and maternal age (p=.027) | *Assumption not met*<br>NCV significant (p=.001)<br>BP significant (BP=22.38, p=.004)<br>Visual inspection of plots – some indication of heteroscedasticity | *Assumption met*<br>Residuals – normally distributed<br>Median residuals = .69<br>Q-Q plot – straight line<br>Shapiro-Wilk not significant (W=.996, p=.647) |
| FD UF-R | *Slight deviations from linearity*<br>Visual inspection of plots suggested no major deviations<br>Tukey's test was significant (p<.001), driven by the Bayley's cognitive score (p<.001) and maternal age (p=.029) | *Assumption not met*<br>NCV significant (p<.001)<br>BP significant (BP=23.99, p=.002)<br>Visual inspection of plots – some indication of heteroscedasticity | *Assumption met.*<br>Residuals – normally distributed<br>Median residuals =.46<br>Q-Q plot – straight line<br>Shapiro-Wilk not significant (W=.996, p=.613) |
| CBCL internalising | | | |
| FD UF-L | *Slight deviations from linearity*<br>Visual inspection of plots suggested no major deviations<br>Tukey's test was significant (p<.001), driven by the Bayley's cognitive score (p=.007). | *Assumption met*<br>NCV not significant (p=.755)<br>BP=1.63, p=.990<br>Visual inspection of plots – ok | *Assumption partially met.*<br>Residuals – relatively normally distributed<br>Median residuals =-.31<br>Q-Q plot – relatively straight line<br>Shapiro-Wilk significant (W=.983, p=.001) |
| FD UF-R | *Slight deviations from linearity*<br>Visual inspection of plots suggested no major deviations<br>Tukey's test was significant (p<.001), driven by the Bayley's cognitive score (p=.007). | *Assumption met*<br>NCV not significant (p=.858)<br>BP=1.45, p=.993<br>Visual inspection of plots - ok | *Assumption partially met.*<br>Residuals – relatively normally distributed<br>Median residuals =.42<br>Q-Q plot – relatively straight line, tails deviating slightly<br>Shapiro-Wilk significant (W=.983, p=.001) |

*FD=fibre density, UF-L=left uncinate fasciculus, UF-R=right uncinate fasciculus, NCV=non-constant variance test, BP=Breusch-Pagan test,*

#### Summary

Visual checks of scatterplots suggested that the relationships between variables are best described by a linear model. However, non-linearity and heteroscedasticity may be driven by influential outliers. These outliers represent accurate values and contain important information, and thus should not be removed from the dataset if this can be avoided. It has been argued that subjective decisions to exclude outliers can be easily abused as a p-hacking strategy (16). Considering the above information, it was determined that robust linear regression would be most appropriate for our data. Robust methods are appropriate to use for real-life datasets, where data distribution is not normal and heavy tails are often present, and they are designed to be resistant to the influence of outliers, non-normality, and model mis-specifications (17)

However, additional sensitivity analyses were conducted removing outliers (e.g. low maternal age, very and very low cognitive scores) and transforming skewed variables. The association between mean FD in the left uncinate fasciculus and total Q-CHAT scores remained significant.

#### Mediation analysis

EPDS scores predicted total Q-CHAT scores (i.e. c path, total effect model) with B=.34, t(307)=3.003, p=.003. In this sample, EPDS did not predict mean FD in the left uncinate fasciculus (i.e. a path, IV predicting mediator), with B=.0003, t(307)=1.734, p=.084. The direct effect model suggested that EPDS (controlling for the mediator, FD UF-L) predicted Q-CHAT scores (i.e. c' path), with B=.31, t(306)=2.720, p=.007), and FD UF-L (controlling for the IV, EPDS) predicted Q-CHAT scores (i.e. b path), with B=101.27, t(306)=3.159, p=.002).

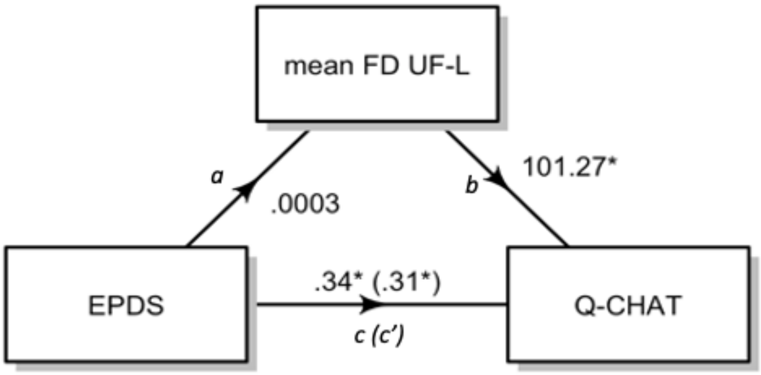


*Fig S5. Diagram for mediation analysis. The path names are provided in italics. B values are presented for each path and * denotes p values significant at p<.05.*

A Sobel test was conducted and found no evidence for mediation in the model (z=1.46, p=.143). Bootstrapped indirect effects also suggested no mediation, with an indirect effect of .035, SE=.02, 95% CI [-.01,.08]. (Fig S5). The analysis was repeated excluding outliers, with similar results.

It is important to note that the mediation analysis was conducted in a smaller sample, and the relationship between EPDS and mean FD in the left uncinate fasciculus was no longer significant in the "a" path.

#### EPDS-3A Exploratory Analysis

In an exploratory analysis, participants were divided into 4 groups based on total EPDS and EPDS-3A scores. EPDS total scores were classed as "high" where participants scored 11 or more, while EPDS-3A scores were classed as "high" where participants scored 6 or more. Scores were as follows: n=32 scored high on both the EPDS and the EPDS-3A, n=350 participants scored low on both, n=19 participants scored high on the total EPDS but low on the EPDS-3A, while n=9 participants scored low on the total EPDS and high on the EPDS-3A.

The sample was too small for any formal analyses, but the distribution of scores for the 4 groups is presented for the mean FD in the left uncinate fasciculus and total Q-CHAT score (Fig S6).

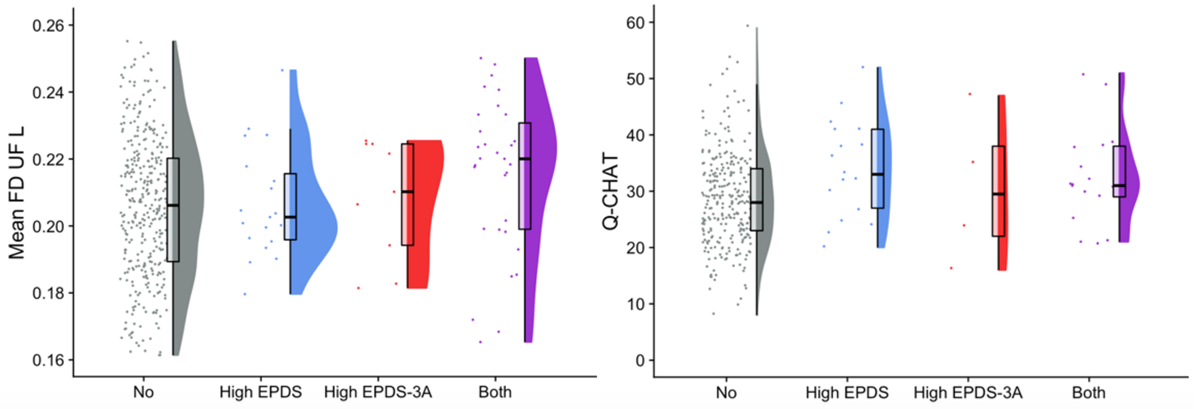


*Fig S6. Raincloud plots showing the distribution of mean FD for the left uncinate fasciculus and Q-CHAT for participants with low total EPDS and EPDS-3A scores ("No"), high total EPDS and low EPDS-3A scores ("High EPDS"), low total EPDS and high EPDS-3A scores ("High EPDS-3A"), and high scores on both the total EPDS and the EPDS-3A ("Both"). For each group, jittered raw data are shown on the left; boxplots with median and interquartile range are shown in the middle; and density plots are shown on the right.*

### Additional figures

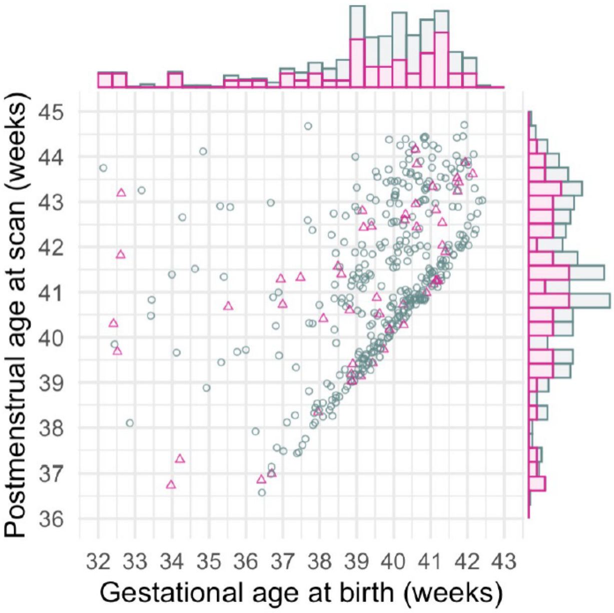


*Fig S7. Age distribution of infants included in the sample; infants born to mothers with high EPDS scores (11 or more) are represented by the pink triangles. Data points are jittered by 0.2 for improved visualisation. The top and right-hand side of the figures contain histograms to summarise the age distribution and using the same colour coding.*

Table S12. Partial (Cohen's) $f^2$ for selected linear regression models, as calculated using the sensemakr package in R (18)

| Model | Intercept | EPDS | Sex | GA at birth | EPDS x Sex | GA at scan | IMD | History of poor mental health | ICV |
|---|---|---|---|---|---|---|---|---|---|
| UF-L FD | .28 | .023 | .043 | .022 | - | .819 | .002 | .002 | - |
| UF-R FD | .30 | .024 | .044 | .024 | - | .785 | .002 | .0009 | - |
| UF-L FDC | .25 | .028 | .016 | .016 | .018 | .217. | .001 | .003 | .355 |
| UF-R FDC | .28 | .013 | .000048 | .019 | - | .232 | .002 | .002 | .352 |
| CD-L FDC | .06 | .011 | .002 | .024 | - | .085 | .003 | .0006 | .320 |

UF-L = left uncinate fasciculus, UF-R = right uncinate fasciculus, CD-L = left dorsal cingulum, FD=fibre density, FDC=fibre density and cross-section, GA=gestational age, IMD=Index of Multiple Deprivation.$f^2 \geq 0.02$, $f^2 \geq 0.15$, and $f^2 \geq 0.35$ represent small, medium, and large effect sizes, respectively. Please note that partial $f^2$ cannot be calculated for the QCHAT models, which were obtained using robust regression.

#### Contributor role taxonomy

The authors contributed to the article as follows: conceptualisation (AL,SV,SJC,MCC,ADE,AB), funding acquisition (ADE), data collection (AC,SF,AL), data curation and analysis (AL,AB,MP,DB,LCG,JDT,DC,JVH), supervision and project administration (AB,SV,MCC,ADE,SJC,CN,JDT,JVH), writing original draft (AL, AB), review and editing (AL,AB,MP,DB,CN,SV,MCC,ADE,SJC,LCG,JDT,DC,JVH,AC,SF). All authors approved the final version of the article prior to submission.
